# Risk of severe COVID-19 outcomes associated with immune-mediated inflammatory diseases and immune modifying therapies: a nationwide cohort study in the OpenSAFELY platform

**DOI:** 10.1101/2021.09.03.21262888

**Authors:** Brian MacKenna, Nicholas A. Kennedy, Amir Mehkar, Anna Rowan, James Galloway, Kathryn E. Mansfield, Katie Bechman, Julian Matthewman, Mark Yates, Jeremy Brown, Anna Schultze, Sam Norton, Alex J. Walker, Caroline E Morton, David Harrison, Krishnan Bhaskaran, Christopher T. Rentsch, Elizabeth Williamson, Richard Croker, Seb Bacon, George Hickman, Tom Ward, Simon Davy, Amelia Green, Louis Fisher, William Hulme, Chris Bates, Helen J. Curtis, John Tazare, Rosalind M. Eggo, David Evans, Peter Inglesby, Jonathan Cockburn, Helen I. McDonald, Laurie A. Tomlinson, Rohini Mathur, Angel YS Wong, Harriet Forbes, John Parry, Frank Hester, Sam Harper, Ian J. Douglas, Liam Smeeth, Charlie W Lees, Stephen JW Evans, Ben Goldacre, Catherine Smith, Sinéad M. Langan

**Affiliations:** The DataLab, Nuffield Department of Primary Care Health Sciences, University of Oxford, OX26GG; Department of Gastroenterology, Royal Devon & Exeter NHS Foundation Trust, Exeter, UK; IBD Research Group, University of Exeter, Exeter, UK; Centre of Rheumatic Diseases, King’s College London, Denmark Hill, London SE5 9RS; London School of Hygiene & Tropical Medicine, Keppel Street, London WC1E 7HT; TPP, TPP House, 129 Low Lane, Horsforth, Leeds, LS18 5PX; Intensive Care National Audit & Research Centre (ICNARC), High Holborn, London WC1V 6AZ; Centre for Genomics and Experimental Medicine, University of Edinburgh Western General Hospital, Edinburgh, UK; St John’s Institute of Dermatology, Guys and St Thomas’ NHS Foundation Trust and Kings College London SE1 9RT

**Author notes:** **Corresponding author:** Professor Sinéad M Langan, Faculty of Epidemiology and Population Health, London School of Hygiene and Tropical Medicine, Keppel Street, WC1E 7HT. Joint first authors (Brian MacKenna, Nicholas A. Kennedy, Amir Mehkar, Anna Rowan, James Galloway). Joint last authors (Stephen JW Evans, Ben Goldacre, Catherine Smith, Sinéad M. Langan).

**Keywords:** immune mediated inflammatory disease, COVID-19 death, COVID-19 critical care admission, COVID-19 hospitalisation, rheumatoid arthritis, psoriatic arthritis, ankylosing spondylitis, inflammatory bowel disease, Crohn’s disease, ulcerative colitis, psoriasis, hidradenitis suppurativa, immune modifying drugs, TNF inhibitor, IL-12/23 inhibitor, I-17 inhibitor, IL-6 inhibitor, JAK inhibitor, rituximab

## Abstract

**Background:** It is unclear if people with immune-mediated inflammatory diseases (IMIDs) (joint, bowel and skin) and on immune modifying therapy have increased risk of serious COVID-19 outcomes.

**Methods:** With the approval of NHS England we conducted a cohort study, using OpenSAFELY, analysingroutinely-collected primary care data linked to hospital admission, death and previously unavailable hospital prescription data. We used Cox regression (adjusting for confounders) to estimate hazard ratios (HR) comparing risk of COVID-19-death, death/critical care admission, and hospitalisation (March to September 2020) in: 1) people with IMIDs compared to the general population; and 2) people with IMIDs on targeted immune modifying drugs (e.g., biologics) compared to standard systemic treatment (e.g., methotrexate).

**Findings:** We identified 17,672,065 adults; of 1,163,438 (7%) with IMIDs, 19,119 people received targeted immune modifying drugs, and 200,813 received standard systemics. We saw evidence of increased COVID-19-death (HR 1.23, 95%CI 1.20, 1.27), and COVID-19 hospitalisation (HR 1.32, 95%CI 1.29, 1.35) in individuals with IMIDs overall compared to individuals without IMIDs of the same age, sex, deprivation and smoking status. We saw no evidence of increased COVID-19 deaths with targeted compared to standard systemic treatments (HR 1.03, 95%CI 0.80, 1.33). There was no evidence of increased COVID-19-related death in those prescribed TNF inhibitors, IL-12/23, IL7, IL-6 or JAK inhibitors compared to standard systemics. Rituximab was associated with increased COVID-19 death (HR 1.68, 95%CI 1.11, 2.56); however, this finding may relate to confounding.

**Interpretation:** COVID-19 death and hospitalisation was higher in people with IMIDs. We saw no increased risk of adverse COVID-19 outcomes in those on most targeted immune modifying drugs for IMIDs compared to standard systemics.

**RESEARCH IN CONTEXT:** *Evidence before this study:* We searched PubMed on May 19^th^, 2021, using the terms “COVID-19”, “SARS-CoV-2” and “rheumatoid arthritis”, “psoriatic arthritis” “ankylosing spondylitis”, “Crohn’s disease” “ulcerative colitis” “hidradenitis suppurativa” and “psoriasis”, to identify primary research articles examining severe COVID-19 outcome risk in individuals with immune-mediated inflammatory diseases (IMIDs) and those on immune modifying therapy. The studies identified (including matched cohort studies and studies in disease-specific registries) were limited by small sample sizes and number of outcomes. Most studies did not show a signal of increased adverse COVID-19 outcomes in those on targeted therapies, with the exception of rituximab. Additionally, disease-specific registries are subject to selection bias and lack denominator populations.

*Added value of the study:* In our large population-based study of 17 million individuals, including 1 million people with IMIDs and just under 200,000 receiving immune modifying medications, we saw evidence that people with IMIDs had an increased risk of COVID-19-related death compared to the general population after adjusting for potential confounders (age, sex, deprivation, smoking status) (HR 1.23, 95%CI 1.20, 1.27). We saw differences by IMID type, with COVID-19-related death being increased by the most in people with inflammatory joint disease (HR 1.47, 95%CI 1.40, 1.54). We also saw some evidence that those with IMIDs were more likely, compared to the general population, to have COVID-19-related critical care admission/death (HR 1.24, 95%CI 1.21, 1.28) and hospitalisation (HR 1.32, 95%CI 1.29, 1.35). Compared to people with IMIDs taking standard systemics, we saw no evidence of differences in severe COVID-19-related outcomes with TNF inhibitors, IL-17 inhibitors, IL-12/23 inhibitors, IL-6 inhibitors and JAK inhibitors. However, there was some evidence that rituximab was associated with an increased risk of COVID-19-related death (HR 1.68, 95%CI 1.11, 2.56) and death/critical care admission (HR 1.92, 95%CI 1.31, 2.81). We also saw evidence of an increase in COVID-19-related hospital admissions in people prescribed rituximab (HR 1.59, 95%CI 1.16, 2.18) or JAK inhibition (HR 1.81, 95%CI 1.09, 3.01) compared to those on standard systemics, although this could be related to worse underlying health rather than the drugs themselves, and numbers of events were small. This is the first study to our knowledge to use high-cost drug data on medicines supplied by hospitals at a national scale in England (to identify targeted therapies). The availability of these data fills an important gap in the medication record of those with more specialist conditions treated by hospitals creating an important opportunity to generate insights to these conditions and these medications

*Implications of all of the available evidence:* Our study offers insights into future risk mitigation strategies and SARS-CoV-2 vaccination priorities for individuals with IMIDs, as it highlights that those with IMIDs and those taking rituximab may be at risk of severe COVID-19 outcomes. Critically, our study does not show a link between most targeted immune modifying medications compared to standard systemics and severe COVID-19 outcomes. However, the increased risk of adverse COVID-19 outcomes that we saw in people with IMIDs and those treated with rituximab merits further study.

## BACKGROUND

Although most people with coronavirus disease 2019 (COVID-19) experience mild symptoms, 15% develop pneumonia requiring hospitalisation, and 5% progress to severe disease (systemic hyperinflammation, major coagulation problems and acute respiratory distress, with risk of multi-organ failure and death) (of unvaccinated individuals).^1^ It is unclear to what extent having immune-mediated inflammatory diseases (IMIDs) – including inflammatory joint (rheumatoid arthritis [RA], psoriatic arthritis [PsA], ankylosing spondylitis [AS]), bowel (Crohn’s disease, ulcerative colitis [UC]), and skin (psoriasis, hidradenitis suppurativa [HS]) diseases – or being on targeted immune modifying therapies for IMID treatment, increases severe COVID-19 outcomes.2

Different strategies have been implemented worldwide to limit infection spread. Policies were initially informed by expert opinion and subsequently by risk estimates for COVID-19 death.3 Early in the pandemic, the UK government issued guidance through the National Health Service (NHS), known as “shielding”, advising individuals considered at high risk of severe COVID-19 to stay at home, minimising face-to-face contacts. Shielding advice targeted individuals identified using health records as having specific immunosuppressive diseases or prescriptions for immune modifying drugs.^4^ Many people with IMIDs taking immune modifying therapies were considered “high risk”, based on prior evidence in other infections.^5,6^

Disease-specific registries have been used to evaluate COVID-19 risks in individuals with IMIDs and those prescribed immune modifying therapies; results have largely been reassuring.^7–9^ However, disease-specific registries are limited by small sample sizes, inherent selection bias, and lack of denominators. OpenSAFELY-TPP is a new secure analytics platform for electronic health records (EHRs) to deliver pandemic-related research.

We aimed to investigate risks of severe COVID-19 outcomes in people with IMIDs and those on targeted immune modifying therapies using English population-based EHR data linked to a new unique national dataset containing information on targeted immune modifying therapies.

## METHODS

### Study design

We conducted a cohort study using electronic health record (EHR) data. We compared risks of COVID-19-related death, critical care admission or death, and hospital admission in: 1) people with IMIDs compared to the general population; and 2) in people with IMIDs prescribed targeted immune modifying drugs compared to those with IMIDs taking standard systemic immune modifying drugs.

### Data source

We used primary care records managed by the GP software provider TPP linked to: Office for National Statistics (ONS) death data, COVID-19 testing data, and a unique national hospital medications dataset (including “high-cost drugs”, typically newer, more expensive medications [e.g., adalimumab] supplied by hospitals for ongoing disease management, **Supplementary Text S1**). We accessed all data through OpenSAFELY, a data analytics platform created by our team for NHS England (https://opensafely.org). OpenSAFELY provides a secure software interface allowing analysis of pseudonymized primary care records in near real-time within the EHR vendor’s highly secure data centre, avoiding the need for data transfer off-site (minimising re-identification risk). Similarly, pseudonymized datasets from other data providers are securely provided by the EHR vendor and linked to primary care data. The dataset analysed within OpenSAFELY was based on 24 million people currently registered with GP surgeries.

### Study population

Our overall study population included adults aged 18–110 years (March 2020), registered with TPP practices with at least 12 months of primary care records prior to March 2020 (**Figure 1**). We followed individuals from March 1^st^, 2020 (UK SARS-Cov-2 outbreak start) until 30^th^ September 2020 (study end), or until the specific outcome under analysis (i.e., COVID-19-related: death, critical care admission or death, or hospitalisation).

**Figure 1.**
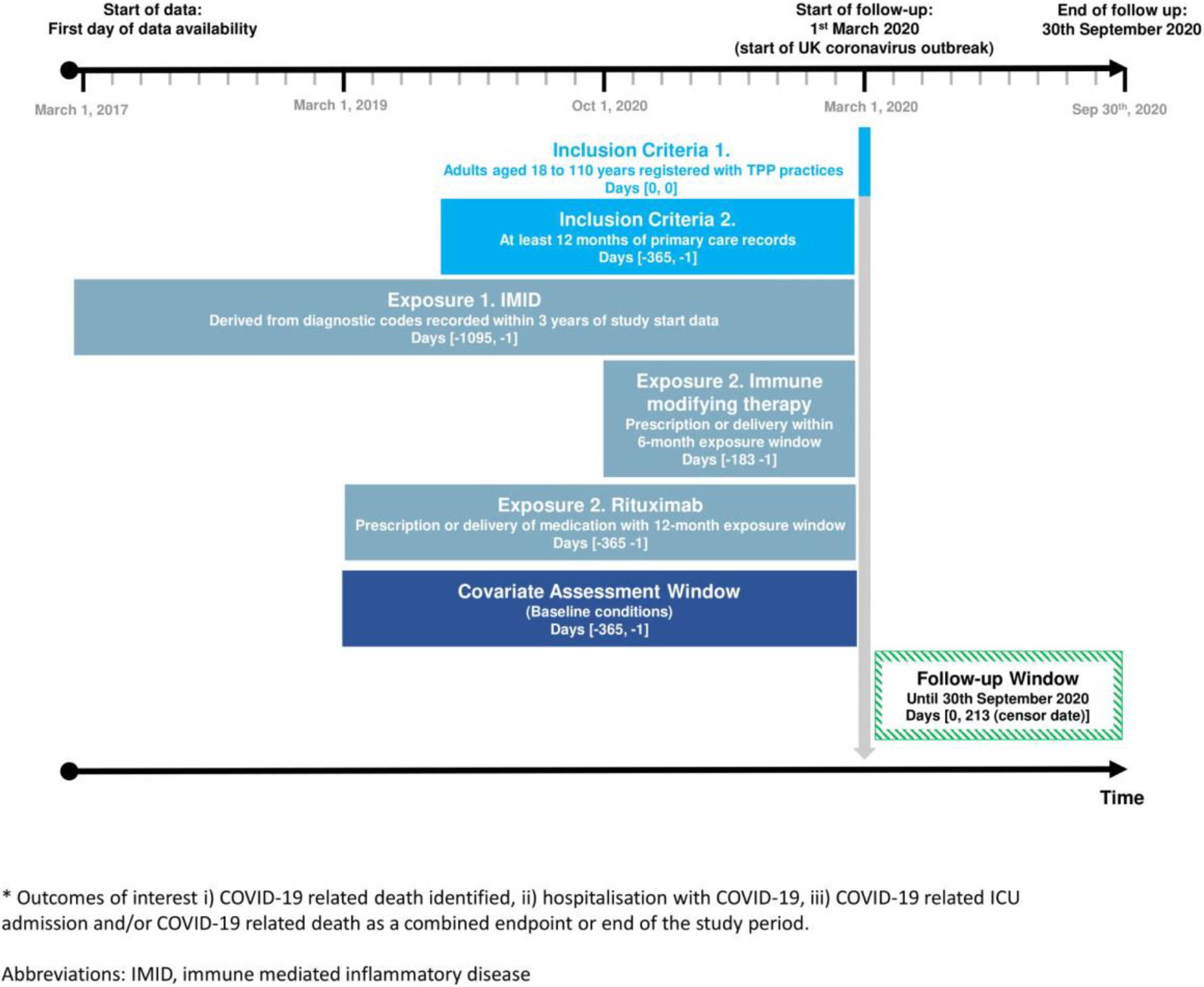
Study design

We compiled diagnostic and therapeutic code lists (in machine readable languages, e.g., SNOMED-CT, NHS dictionary of medicines and devices) for all study variables (exposures, outcomes, covariates). Detailed information on compilation and sources of code lists are freely available for inspection and re-use (https://codelists.opensafely.org/).

### Exposures

Exposures were: 1) IMIDs: inflammatory joint disease (RA, PsA, AS), inflammatory bowel disease (IBD) (Crohn’s disease, UC, IBD unclassified), and inflammatory skin disease (psoriasis, HS); and 2) prescription of systemic immune modifying medication by general practitioners or supplied by hospitals through high-cost drugs procedures.

We identified people with IMIDs using diagnostic morbidity codes in primary care during the three years prior to March 1^st^, 2020. People with IMID diagnoses in multiple categories (e.g., inflammatory joint and skin disease) contributed to comparisons with the general population for all IMID categories they had records for (e.g., individuals with PsA and psoriasis, contributed to both joint and skin disease).

Immune modifying medications were categorised as standard systemic therapy and targeted therapy. Standard systemic therapies included leflunomide, methotrexate, mycophenolate mofetil or mycophenolic acid, ciclosporin, sulphasalazine, mercaptopurine, thioguanine, and azathioprine. Targeted therapies included biologic therapies: TNF inhibitors (etanercept, adalimumab, golimumab, certolizumab, infliximab, and biosimilar versions of these medications), IL-17 inhibitors (secukinumab, ixekizumab, brodalumab), IL-12/23 inhibitors (ustekinumab, guselkumab, risankizumab, tildrakizumab), IL-6 inhibitors (tocilizumab, sarilumab), B-cell depletion therapy (rituximab), and novel small molecules: Janus Kinase (JAK) inhibitors (baricitinib, tofacitinib).^10–15^ Individuals treated with both systemic therapy and targeted therapies were considered exposed to targeted therapies.

We identified standard systemic therapies using primary care prescribing data. We identified targeted immune modifying medications using high-cost drugs invoicing **(Supplementary Text S1)**. Drug exposure was defined by at least one prescription, or delivery of medication to an individual before March 1^st^, 2020 (date chosen as some of medications were either specifically used, or stopped, due to the pandemic). For each individual, we defined drug exposure based on the closest drug recorded prior to the study start (March 1^st^ 2020) allowing for a maximum of 6 months before the start of the study for all agents apart from rituximab, where we permitted a 12-month exposure window (to allow for the frequency of treatment and the longer duration of response of rituximab, including known prolonged effects on vaccine responses).^16,17^ We could not evaluate medication switching or adherence during the follow-up period, as data were only available up to March 2020.

### Outcomes

Outcomes were COVID-19-related death, critical care admission or death, and hospitalisation. We identified COVID-19-related deaths based on records of COVID-related ICD-10 codes (U071, U072) anywhere on death certificates. We used COVID-19-related critical care admission (Intensive Care National Audit & Research Centre^18^) or death as a combined endpoint to reflect individuals with severe COVID-19 who died without being admitted to a critical care unit for critical care admission. We identified COVID-19-related hospitalisation as a positive polymerase chain reaction (PCR) test less than 28 days before admission and, up to 5 days post admission to exclude nosocomial infection.

### Covariates

We selected potential confounders and mediators *a priori* based on clinical knowledge and prior evidence.^2^ In the relationship between IMIDs and severe COVID-19 outcomes, we considered age (categorical variable), sex, deprivation (using quintiles of Index of Multiple Deprivation),^19,20^ and smoking status to be potential confounders; we considered BMI, cardiovascular disease, diabetes and current glucocorticoid use to be potential mediators. In the relationship between (choice of) immune modifying therapy and severe COVID-19 outcomes we considered age, sex, deprivation, smoking status, BMI, specific IMID (inflammatory joint, bowel and skin disease), cardiovascular disease, cancer (excluding non-melanoma skin cancer), stroke, end stage renal failure, chronic liver disease, chronic respiratory disease and diabetes mellitus as potential confounders; we considered current glucocorticoid use as a potential mediator. Details of covariate definitions are included in **Supplementary Text S2**. Figures representing assumed relationships between covariates, primary exposures and outcomes are in **Supplementary Figures S1 and S2.**

### Statistical methods

We initially described characteristics of the general population, people with IMIDs, and those with IMIDs prescribed immune modifying therapy. We used Cox regression to estimate hazard ratios (HR) (95%CI), comparing people with IMIDs to the general population, and people with IMIDs on standard systemic drugs to those on targeted therapies. We adjusted models for confounding based on assumptions inherent in our conceptual frameworks (**Supplementary Figures S1, S2**). For comparisons between those with IMIDs to the general population, we adjusted for age, sex, deprivation, and smoking. For comparisons between those on targeted therapies to those on standard systemic drugs, we adjusted for age, sex, deprivation, obesity (BMI>=30), smoking, IMIDs (bowel, joint, skin), and comorbidities (cardiovascular disease, diabetes mellitus, cancer, stroke, end stage renal failure, chronic liver disease, chronic respiratory disease). We also fitted models adjusted for additional covariates considered to be mediators. We tested Cox model assumptions using Schoenfeld residuals.

#### Sensitivity analyses

We repeated our main analyses in sensitivity analyses assessing robustness of our findings (**Supplementary Table S1, Text S3**). We considered immune-mediated inflammatory disease severity and degree of shielding to be potential unmeasured confounders of associations between specific immune-modifying therapy and COVID-19 outcomes. We therefore conducted quantitative bias analysis using E-values to assess how strongly associated unmeasured confounders would need to be with exposure and outcome to potentially fully explain observed non-null associations (i.e., association adjusted for both measured covariates and the unmeasured confounder would be null).^21^

### Software and reproducibility

We used Python for data management, and Stata 16/Python for analyses. All code used for data management and analyses is available online (https://github.com/opensafely/immunosuppressant-meds-research) including all iterations of the pre-specified study protocol archived with version control.

### Patient and public involvement

Patients were not formally involved in developing this study design, which was developed rapidly during a pandemic. We invite patients or members of the public to contact us through our website https://opensafely.org/ regarding this study or the broader OpenSAFELY project. Our protocol and paper preprint have been sent to Crohn’s and Colitis UK, Psoriasis Association, National Rheumatoid Arthritis Society, and Versus Arthritis for review from an expert-patient perspective.

### Ethics

The study was approved by the Health Research Authority (Research Ethics Committee reference 20/LO/0651) and the London School of Hygiene and Tropical Medicine’s Ethics Board (Reference 21863).

## RESULTS

Of 17,672,065 people in the overall study population (**Supplementary Figure S3**), 1,163,438 (7%) had an IMID diagnosis (**Table 1**). Of those with IMIDs: 1) 23% (n=272,452) had inflammatory joint disease (RA 16% n=183,485, PsA 5% n=54,593, AS 3% n=35,138); 2) 17% (n=199,037) had IBD (Crohn’s 35% n=69,788, UC 51% n=100,617, unclassified IBD 16% n=32,093); and 3) 66% (n=769,816) had inflammatory skin disease (psoriasis 90% n=693,178, HS 10% n=76,746).

**Table 1:** Descriptive Characteristics of general population and people with IMIDs.

|  | General population | Overall IMID population | Inflammatory joint disease | Inflammatory skin disease | Inflammatory bowel disease |
| --- | --- | --- | --- | --- | --- |
|  | N=16,508,627 | N=1,163,438 | N=272,452 | N=769,816 | N=199,037 |
| **Age** |  |  |  |  |  |
| 18-39 | 5,808,217 (35.2%) | 252,718 (21.7%) | 25,238 (9.3%) | 191,634 (24.9%) | 46,099 (23.2%) |
| 40-49 | 2,727,833 (16.5%) | 183,130 (15.7%) | 32,366 (11.9%) | 130,758 (17.0%) | 32,057 (16.1%) |
| 50-59 | 2,882,387 (17.5%) | 232,525 (20.0%) | 56,192 (20.6%) | 155,223 (20.2%) | 39,513 (19.9%) |
| 60-69 | 2,235,982 (13.5%) | 209,384 (18.0%) | 62,359 (22.9%) | 129,432 (16.8%) | 34,853 (17.5%) |
| 70-79 | 1,797,487 (10.9%) | 186,613 (16.0%) | 62,200 (22.8%) | 107,331 (13.9%) | 31,215 (15.7%) |
| 80+ | 1,056,721 (6.4%) | 99,068 (8.5%) | 34,097 (12.5%) | 55,438 (7.2%) | 15,300 (7.7%) |
| **Sex** |  |  |  |  |  |
| Male | 8,293,607 (50.2%) | 523,274 (45.0%) | 107,104 (39.3%) | 356,220 (46.3%) | 96,054 (48.3%) |
| **BMI** |  |  |  |  |  |
| Underweight (<18.5) | 314,887 (1.9%) | 21,231 (1.8%) | 5,995 (2.2%) | 11,280 (1.5%) | 5,158 (2.6%) |
| Normal (18.5-24.9) | 4,576,346 (27.7%) | 306,029 (26.3%) | 74,283 (27.3%) | 186,383 (24.2%) | 63,902 (32.1%) |
| Overweight (25.0-29.9) | 4,462,587 (27.0%) | 351,450 (30.2%) | 87,569 (32.1%) | 226,580 (29.4%) | 62,068 (31.2%) |
| Obese I (30.0-34.9) | 2,255,908 (13.7%) | 202,825 (17.4%) | 50,614 (18.6%) | 137,770 (17.9%) | 30,048 (15.1%) |
| Obese II (35.0-39.9) | 871,125 (5.3%) | 88,344 (7.6%) | 21,818 (8.0%) | 62,536 (8.1%) | 11,135 (5.6%) |
| Obese III (40.0+) | 502,285 (3.0%) | 55,834 (4.8%) | 12,896 (4.7%) | 41,747 (5.4%) | 5,744 (2.9%) |
| Missing | 3,525,489 (21.4%) | 137,725 (11.8%) | 19,277 (7.1%) | 103,520 (13.4%) | 20,982 (10.5%) |
| **Index of multiple deprivation** |  |  |  |  |  |
| 1 (least deprived) | 3,337,475 (20.2%) | 242,175 (21.8%) | 57,464 (21.1%) | 156,444 (20.3%) | 44,874 (22.5%) |
| 2 | 3,280,436 (19.9%) | 235,706 (20.3%) | 56,059 (20.6%) | 152,956 (19.9%) | 42,621 (21.4%) |
| 3 | 3,294,811 (20.0%) | 233,866 (20.1%) | 56,398 (20.7%) | 152,627 (19.8%) | 40,775 (20.5%) |
| 4 | 3,330,769 (20.2%) | 228,552 (19.6%) | 53,089 (19.5%) | 152,678 (19.8%) | 37,674 (18.9%) |
| 5 (most deprived) | 3,129,886 (19.0%) | 213,903 (18.4%) | 47,616 (17.4%) | 148,866 (19.3%) | 31,274 (15.7%) |
| Missing | 135,250 (0.8%) | 9,236 (0.8%) | 1,826 (0.7%) | 6,245 (0.8%) | 1,819 (0.9%) |
| **Smoking** |  |  |  |  |  |
| Never | 7,687,903 (46.6%) | 420,806 (36.2%) | 102,798 (37.7%) | 265,169 (34.4%) | 79,651 (40.0%) |
| Former | 5,310,393 (32.2%) | 509,886 (43.8%) | 128,484 (47.2%) | 327,811 (42.6%) | 91,893 (46.2%) |
| Current | 2,774,203 (16.8%) | 220,916 (19.0%) | 40,007 (14.7%) | 168,056 (22.8%) | 25,350 (12.7%) |
| Missing | 736,1280 (4.5%) | 11,8300 (1.0%) | 1,163 (0.4%) | 8,780 (1.1%) | (2,143 (1.1%) |
| **Comorbidities** |  |  |  |  |  |
| *Diabetes mellitus* |  |  |  |  |  |
| With HbA1c<58mmol/mol | 1,033,685 (6.3%) | 112,193 (9.6%) | 32,631 (12.0%) | 71,520 (9.3%) | 17,366 (8.7%) |
| With HbA1c≥58mmol/mol | 456,388 (2.8%) | 48,951 (4.2%) | 13,058 (4.8%) | 32,388 (4.2%) | 7,766 (3.9%) |
| With unknown HbA1c | 240,398 (1.5%) | 21,567 (1.9%) | 5,741 (2.1%) | 14,071 (1.8%) | 3,482 (1.7%) |
| *Cardiovascular disease* | 1,146,032 (6.9%) | 129,065 (11.1%) | 42,078 (15.4%) | 76,916 (10.0%) | 20,536 (10.3%) |
| *Stroke* | 372,332 (2.3%) | 40,523 (3.5%) | 12,872 (4.7%) | 24,075 (3.1%) | 6,587 (3.3%) |
| *Cancer* | 962,622 (5.8%) | 94,832 (8.2%) | 27,779 (10.2%) | 56,751 (7.4%) | 17,150 (8.6%) |
| *End stage renal failure* | 22,408 (0.1%) | 2,190 (0.2%) | 580 (0.2%) | 1,217 (0.2%) | 550 (0.3%) |
| *Chronic respiratory disease* | 666,384 (4.0%) | 94,350 (8.1%) | 33,690 (12.4%) | 53,614 (7.0%) | 14,725 (7.4%) |
| *Chronic liver disease* | 98,012 (0.6%) | 15,333 (1.3%) | 3,877 (1.4%) | 9,340 (1.2%) | 3,758 (1.9%) |
| **Glucocorticoid use** |  |  |  |  |  |
| 1 or more prescription in last 3 months* | 317,938 (1.9%) | 64,151 (5.5%) | 30,928 (11.4%) | 27,673 (3.6%) | 11,913 (6.0%) |
| **Ethnicity\*\*** |  |  |  |  |  |
| White | 10,614,096 (64.3%) | 827,457 (71.1%) | 195,851 (71.9%) | 547,080 (71.1%) | 141,986 (71.3%) |
| Asian /Asian British | 999,881 (6.1%) | 50,382 (4.3%) | 12,771 (4.7%) | 31,964 (4.2%) | 8,685 (4.4%) |
| Black | 340,723 (2.1%) | 9,960 (0.9%) | 2,723 (1.0%) | 6,071 (0.8%) | 1,502 (0.8%) |
| Mixed/other | 494,119 (3.0%) | 16,797 (1.4%) | 3,655 (1.3%) | 11,175 (1.5%) | 2,736 (1.4%) |
| Missing | 4,059,808 (24.6%) | 258,842 (22.2%) | 57,452 (21.1%) | 173,526 (22.5%) | 44,128 (22.2%) |

All figures are n (%) unless otherwise specified.
People with diagnoses across subcategories contributed to multiple categories (e.g., someone with psoriasis and PsA, contributed to both skin and joint categories of IMID), therefore individuals may be included in more than one IMID category.
\*Glucocorticoid use refers to individuals with one or more prescriptions for any dose of oral glucocorticoid in the last 3 months prior to study start.
\*\* Ethnicity data are included for interest. However, ethnicity was not adjusted for in the main analysis due to the high proportion of missing data, although we did adjust for ethnicity in a sensitivity analysis (**Supplementary Table S3**).
**Abbreviations:** IMID, immune mediated inflammatory disease; BMI, Body mass index; HbA1c: glycosylated haemoglobin.

### IMIDs compared to the general population

Compared to the general population, people with IMIDs were older (>=70 years: 25% versus 17% in the general population), more likely to be female (55% versus 50%), of white ethnicity (71% versus 64%), obese (BMI>=30, 30% versus 22%), and with more comorbidities (e.g., cardiovascular disease: 11% versus 7%: diabetes mellitus: 16% versus 11%) (**Table 1**). There were differences between individuals with inflammatory joint, skin and bowel diseases, for example, individuals with inflammatory joint disease were older compared to those with IBD or inflammatory skin disease (>=70 years: 35.3% joint, 23.4% IBD, 21.1% skin).

#### COVID-19-related death

After adjusting for age and sex, compared to the general population, people with IMIDs had greater risk of COVID-19-related death (HR 1.27, 95%CI 1.23, 1.31). Evidence of association between IMIDs and COVID-19-related death remained after additionally adjusting for confounders (age, sex, deprivation and smoking status, HR 1.23, 95%CI 1.20, 1.27) and after further adjusting for potential mediators (BMI, cardiovascular disease, diabetes and current glucocorticoid use, HR 1.15, 95%CI 1.11, 1.18). (**Figure 2, Supplementary Table S2**).

**Figure 2.**
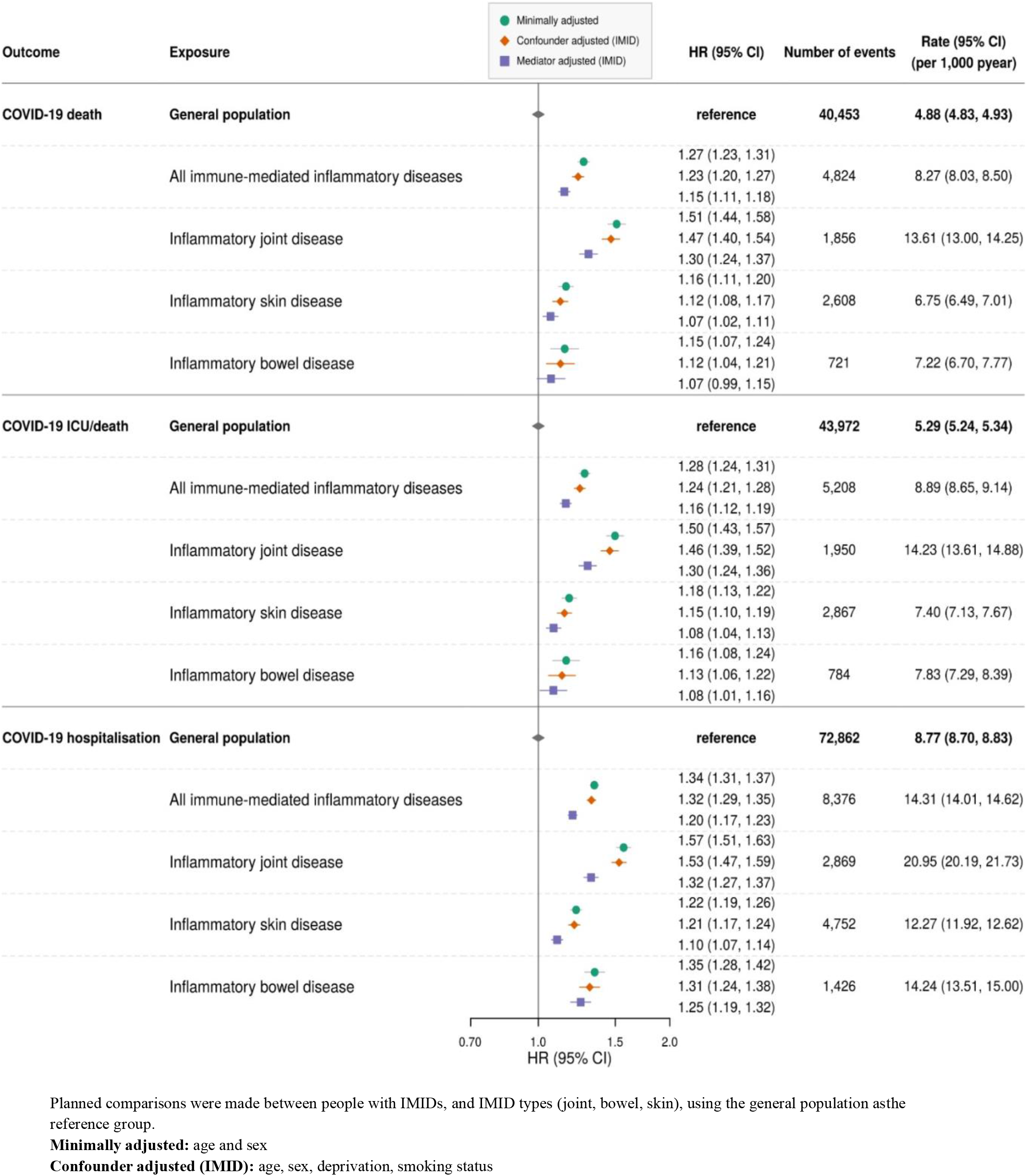
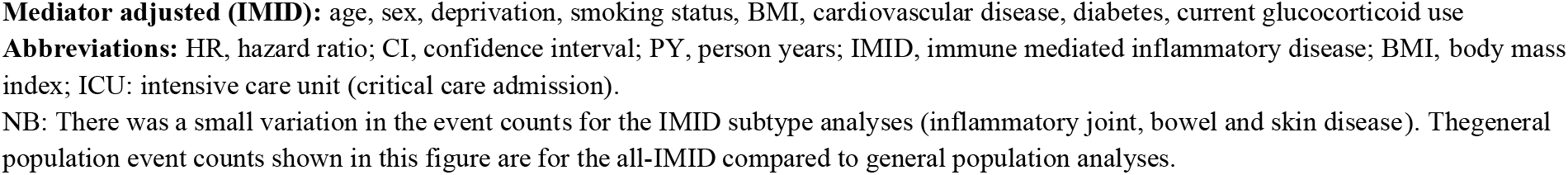
Forest plot of hazard ratios (HRs) for COVID-19-related death, critical care admission/death and hospitalisation for IMID vs general population

After adjusting for age and sex, compared to the general population we saw increased COVID-19-related death in people with inflammatory joint (HR 1.51, 95%CI 1.44, 1.58), bowel (HR 1.15,95%CI 1.07, 1.24), and skin (HR 1.16, 95%CI 1.11, 1.20) diseases. After further adjusting for potential confounders, evidence for association between specific IMID types and COVID-19 related death persisted for all IMID types and was greatest for inflammatory joint disease (HR 1.47, 95%CI 1.40, 1.54), with smaller effect estimates for inflammatory skin (HR 1.12, 95%CI 1.08, 1.17) or bowel (HR 1.12, 95%CI 1.04, 1.21) disease, and further attenuation after adjusting for potential mediators.

#### COVID-19-related critical care admission or death

Individuals with IMIDs had greater risk of COVID-19-related critical care admission or death compared to the general population, which persisted after adjusting for confounders (HR 1.24, 95%CI 1.21, 1.28) and further adjusting for mediators (HR 1.16, 95%CI 1.12, 1.19). Compared to the general population, there was evidence of increased COVID-19-related critical care admission or death in people with inflammatory joint (confounder adjusted HR 1.46, 95%CI 1.39, 1.52), skin (confounder adjusted HR 1.15, 95%CI 1.10, 1.19), and bowel (confounder adjusted HR 1.13, 95%CI 1.06, 1.22) diseases.

#### COVID-19-related hospitalisation

Compared to the general population, people with IMIDs had greater risk of COVID-19-related hospitalisation, which remained after adjusting for potential confounders (HR 1.32, 95%CI 1.29, 1.35) and mediators (HR 1.20, 95%CI 1.17, 1.23). Compared to the general population, risk of COVID-19 hospitalisation was increased in all IMID categories including inflammatory joint (confounder adjusted HR 1.53, 95%CI 1.47, 1.59), skin (adjusted HR 1.21, 95%CI 1.17, 1.24), and bowel (adjusted HR 1.31, 95%CI 1.24, 1.38) disease.

#### Sensitivity analyses

Results from sensitivity analyses were broadly similar to the main analysis. (**Supplementary Tables S3-S4**).

### Targeted therapy compared to standard systemic therapy

Of those with IMIDs, 200,813 individuals (17%) were prescribed either standard systemic therapy (90%, n=181,694/200,813) or targeted immune modifying therapy (10%, n=19,119/200,813) (**Table 2, Supplementary Table S5**). Compared to those on standard systemic therapy, individuals receiving targeted therapy were younger (<40 years: 22% versus 14%) and less likely to have comorbidities (e.g., cardiovascular disease). The most commonly prescribed targeted therapies were TNF inhibitors (71%, n=13,524/19,119), followed by rituximab (10%, n=1,998/19,119); IL-12/23 inhibitors, (7%, n=1,379/19,119); IL-17 inhibitors (5%, n=1,036/19,119); JAK inhibitors (5%, n=871/19,119) and IL-6 inhibitors (4%, n=758/19,119).

**Table 2:**
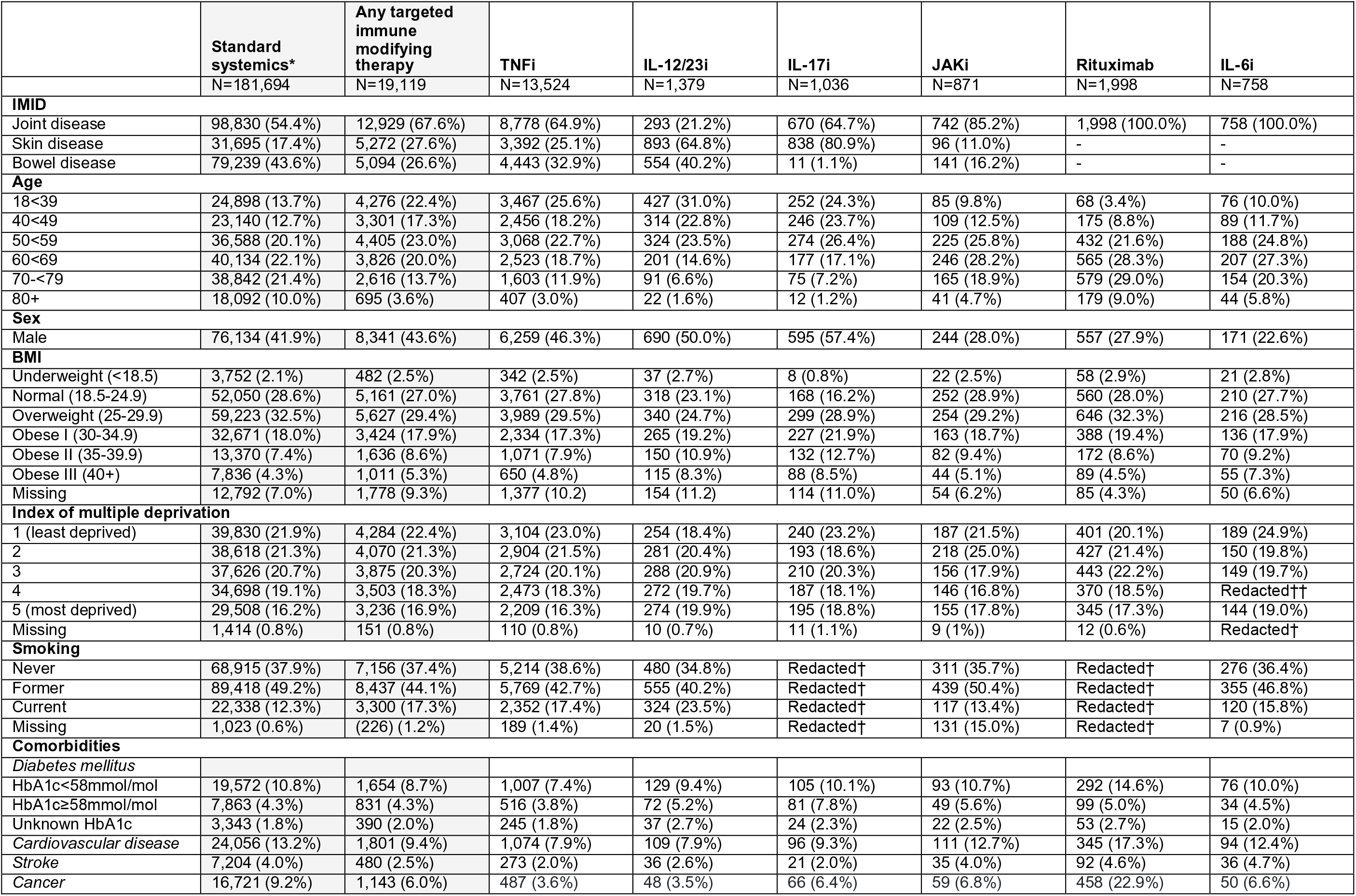
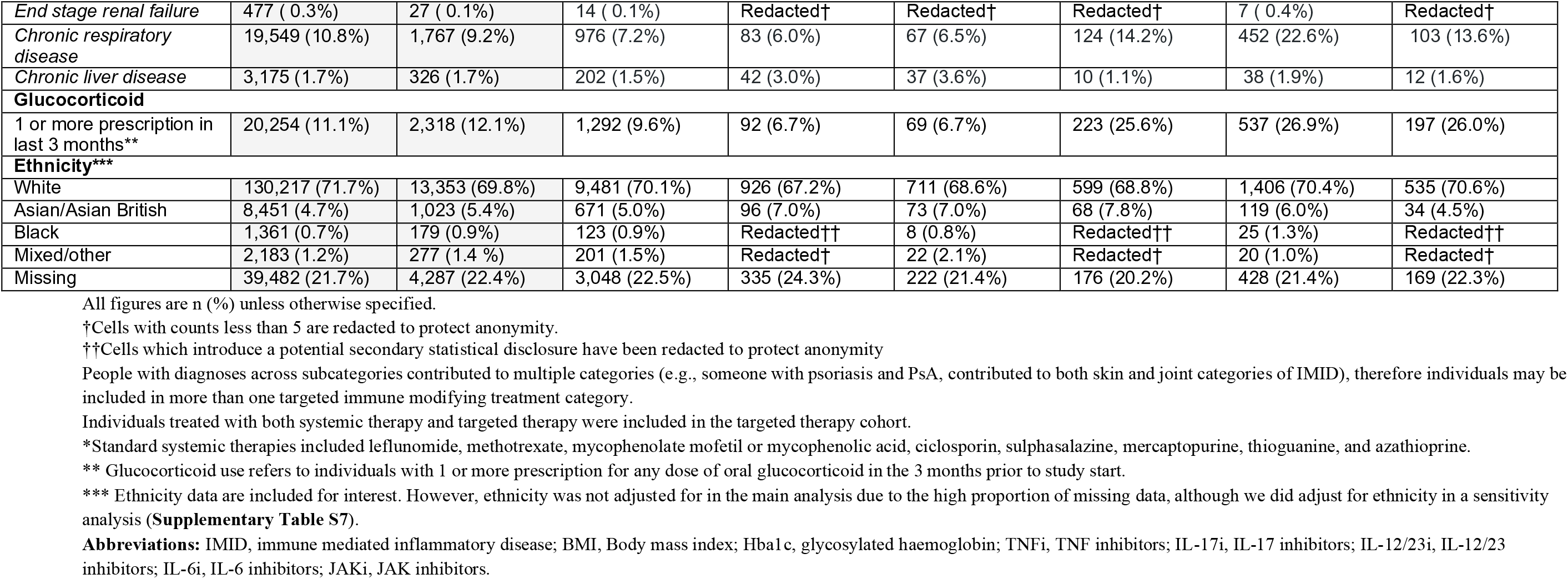
Descriptive characteristics of IMID population on targeted and standard systemic immune modifying therapy.

#### COVID-19-related death

After adjusting for potential confounders (age, sex, deprivation, BMI, IMIDs [bowel, joint, skin], cardiovascular disease, cancer, stroke, diabetes mellitus), we saw no evidence of differences in COVID-19-related death in individuals on targeted therapy overall compared to individuals on standard systemic therapy (HR 1.03, 95%CI 0.80,1.33) (**Figure 3, Supplementary Table S6**).

**Figure 3.**
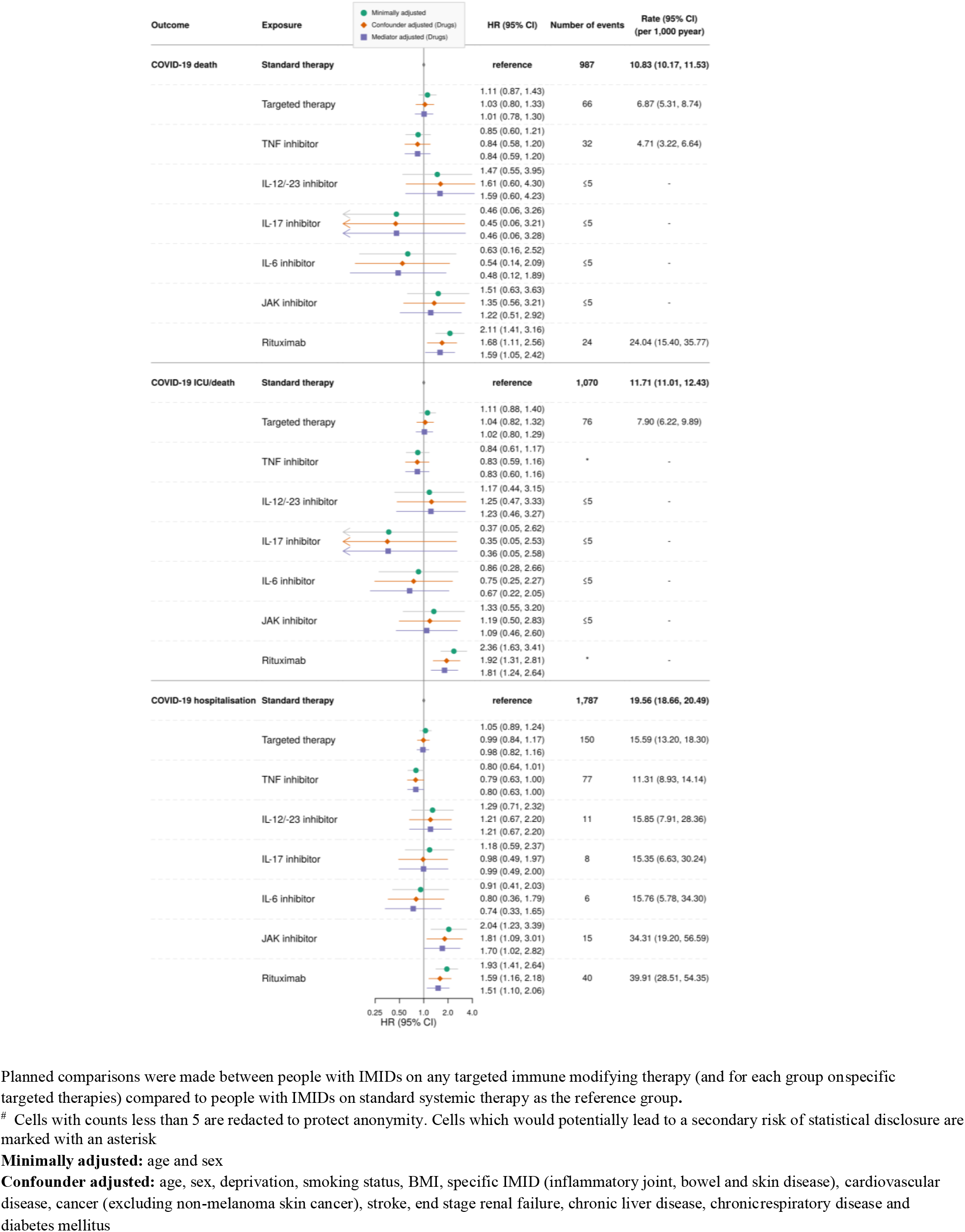
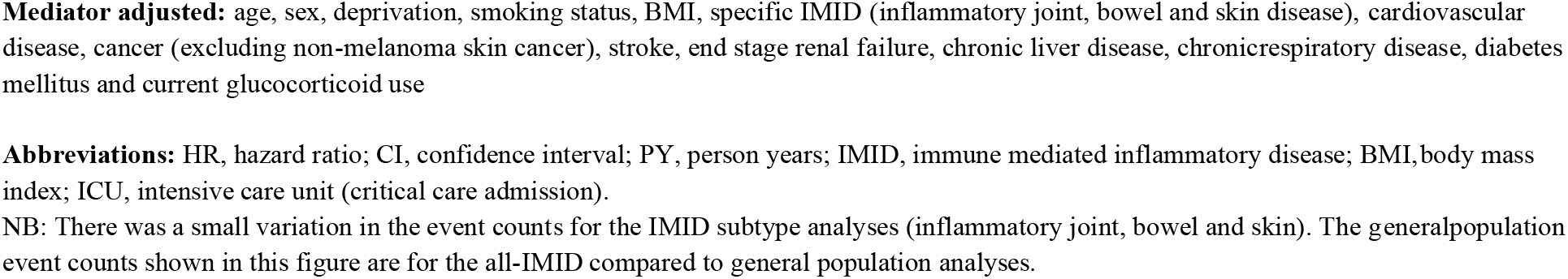
Forest plot of hazard ratios (HRs) for COVID-19 death, critical care admission/death and hospitalisation for standard systemic vs targeted immunosuppression

Compared to individuals on standard systemic therapy, there was no observed increased risk of COVID-19 related death in individuals on TNF inhibitors, IL-12/23 inhibitors, IL7 inhibitors, JAK inhibitors or IL-6 inhibitors (although confidence intervals were wide). Compared to individuals on standard systemic therapy, people receiving rituximab had an increased risk of COVID-19-related death (HR 1.68, 95%CI 1.11, 2.56), based on 24 deaths in the rituximab group.

#### COVID-19-related critical care admission or death

After adjusting for confounders, compared to individuals on standard systemic therapy, there was no increase in COVID-19-related critical care admission or death in individuals on TNF inhibitors, IL-12/23 inhibitors, IL-17 inhibitors, JAK inhibitors or IL-6 inhibitors. People receiving rituximab had increased risk of critical care admission or death (HR 1.92, 95%CI 1.31, 2.81), based on 29 events in the rituximab group.

#### COVID-19-related hospitalisation

After adjusting for confounders, compared to individuals on standard systemic therapy, there was no difference in risk of COVID-19-related hospitalisations in individuals on TNF inhibitors, IL-12/23 inhibitors, IL-17 inhibitors, or IL-6 inhibitors. Compared to standard systemics, we saw increased risk of COVID-19-related hospitalisation in people receiving rituximab (HR 1.59, 95%CI 1.16, 2.18; 40 events), and JAK inhibitors (HR 1.81, 95%CI 1.09, 3.01; 15 events).

#### Sensitivity analyses

Excluding people with haematological cancers and organ transplants attenuated the effect estimate for rituximab (HR 1.54, 95%CI 0.95, 2.49; 18 events). Otherwise, results from sensitivity analyses were similar to the main analysis (**Supplementary Tables S7-S11**). In quantitative bias analysis comparing individuals with IMIDs taking rituximab or JAK inhibitors to standard systemics we note an unmeasured confounder moderately associated with both exposure and outcome could potentially explain associations of rituximab (by at least a risk ratio of 1.68) and JAK inhibitors (by at least a risk ratio of 1.81) with adverse COVID-19 outcomes.

## DISCUSSION

In this large population-based study using data fromOpenSAFELY, we found that people with IMIDs experience higher rates of COVID-19-related death, critical care admission and death, and hospitalisation than people without IMIDs of the same age, sex, deprivation and smoking status. We demonstrated that compared to standard systemic immune modifying therapies for IMIDs, there was no increased risk of COVID-19-related death in those prescribed TNF, IL-12/23, IL7, IL-6 or JAK inhibitors. However, rituximab was associated with an increased risk of death (HR 1.64, 95%CI 1.08, 2.49) and critical care admission, which may relate to confounding.

### Findings in context

Our findings — suggesting that people with IMIDs were at increased risk of COVID-19-related death compared to people without IMIDs of the same age, sex, deprivation, and smoking status — have not previously been reported. This study provides granular data on a range of IMIDs but is consistent with previous less detailed analyses.^22^ Our finding that those with IMIDs were more likely to be hospitalised for COVID-19 compared to the general population is consistent with Canadian and Danish cohort studies ^232423^ and reports of adverse COVID-19 outcomes for people with specific IMIDs.^25^ However, factors leading to adverse COVID-19 outcomes are likely multifactorial, with possible explanations including more severe symptoms following COVID-19, unmeasured confounders, better access to care, or a lower physician threshold for admission in those on immune modifying drugs.

There are no comparable population-based data sources including information on COVID-19 outcomes in those on targeted therapies for IMIDs. Our observation that those on targeted therapies do not have an increased risk of COVID-19 related death is consistent with data from international registries.^7–9^ Compared to our findings, a recent meta-analysis using data from registries included 2,766 individuals with autoimmune diseases and COVID-19 diagnoses, reported higher rates of hospitalisation and death in people prescribed combination standard systemic therapy and biologics or JAK inhibitors, but lower rates in those prescribed TNF inhibitor monotherapy.^26^ The Global Rheumatology Alliance reported no increase in COVID-19-related death with biologic therapies or JAK inhibitors compared to methotrexate monotherapy, but an increase in COVID-19-related death with rituximab.^8^ The Surveillance Epidemiology of Coronavirus Under Research Exclusion for Inflammatory Bowel Disease (SECURE-IBD) reported no association in COVID-19-related death, critical care or hospitalisation in people on TNF inhibitors compared to those not prescribed TNF inhibitor therapy.7

Our observed increased risk of critical care admission or death in people treated with rituximab is consistent with previous reports of increased mortality in people treated with B cell depleting agents (e.g., rituximab for oncology indications).^27^ Explanations could include mechanistic roles of B cells in COVID-19 immune responses, unmeasured characteristics of the RA population treated with rituximab or confounding by severity (i.e., rituximab is preferentially prescribed for seropositive disease, with extra-articular manifestations including pulmonary fibrosis) or by cancer.^16^ We were underpowered to assess effects of regular tocilizumab on COVID-19 outcomes, although trial data has shown benefit in hospitalised and critically ill patients.^28,29^

### Strengths and weaknesses

The key strengths of this study are the scale and completeness of underlying raw EHR data. The OpenSAFELY platform accesses an unprecedented scale of data; the full dataset of all raw, single-event-level clinical events for all individuals at 40% of all GP practices in England, including all tests, treatments, diagnoses, and clinical and demographic information linked to various sources of hospital data including, for the first time, a comprehensive dataset of medications supplied by hospitals. We recognise some limitations. Although English primary care records are longitudinal and comprehensive, certain confounders were not captured. Shielding, as recommended for groups of clinically vulnerable people by the Chief Medical Officer,^30^ may have reduced the risk of infection, thus likely biasing results towards the null. In mediator adjusted models, we adjusted for concomitant use of oral glucocorticoids, however this adjustment is likely to be imperfect, leading to residual confounding. We also considered cardiovascular disease and diabetes mellitus to be mediators in the relationship between IMIDs and severe COVID-19 outcomes, however the timing of mediator assessment at index means they could have predated the IMID diagnosis, and hence not be true mediators. Assessment of glucocorticoid doses and usage is challenging due to absent precise dose information, prescribing of reducing dose regimens, low-cost medication administered in hospital alongside high-cost drugs, pandemic stockpiling and patient-led discontinuation due to COVID-19-related concerns (may also apply to immune modifying medication).

Finally, as with any large study using routinely-collected data, there is a possibility of misclassification of exposure status. Misclassification of high cost drug exposure is unlikely, as high cost drug information is critical for billing. However, it is possible that we may have systematically excluded some individuals taking standard systemic drugs from analyses comparing targeted to standard systemic therapies. These exclusions may result in underestimation of risks in the standard systemic group due to differential exclusion of “new starters”, where the first prescription was in hospital. In practice, we expect the effects of this misclassification to be minimal due to the short time window.

### Implications and summary

We have used one of the largest population-based datasets globally with linked data on immune modifying drugs to describe COVID-19 risks for people with IMIDs. We found that COVID-19 death and hospitalisation were higher in people with IMIDs; however, we saw no increased risk of adverse COVID-19 outcomes in those on most targeted immune modifying drugs for IMIDs compared to standard systemics. The availability of the “high-cost” novel data fills an important gap in available UK EHR data, and routine availability of these data is essential for future research.

Our findings provide an evidence-base to inform policy on booster vaccination prioritisation and risk mitigating behaviour advice, but must be interpreted in the context of UK public health policy on shielding. Findings will support health care professionals engaging in shared decision making and communication of risk.

## Data Availability

NHS England is the data controller; TPP is the data processor; and the key researchers on OpenSAFELY are acting on behalf of NHS England. This implementation of OpenSAFELY is hosted within the TPP environment which is accredited, the ISO 27001 information security standard and is NHS IG Toolkit compliant; patient data has been pseudonymised for analysis and linkage using industry standard cryptographic hashing techniques; all pseudonymised datasets transmitted for linkage onto OpenSAFELY are encrypted; access, the platform is via a virtual private network (VPN) connection, restricted, a small group of researchers; the researchers hold contracts with NHS England and only access the platform, initiate database queries and statistical models; all database activity is logged; only aggregate statistical outputs leave the platform environment following best practice for anonymisation of results such as statistical disclosure control for low cell counts. The OpenSAFELY research platform adheres, the obligations of the UK General Data Protection Regulation (GDPR) and the Data Protection Act 2018. In March 2020, the Secretary of State for Health and Social Care used powers under the UK Health Service (Control of Patient Information) Regulations 2002 (COPI), require organisations, process confidential patient information for the purposes of protecting public health, providing healthcare services, the public and monitoring and managing the COVID-19 outbreak and incidents of exposure; this sets aside the requirement for patient consent. Taken together, these provide the legal bases, link patient datasets on the OpenSAFELY platform. GP practices, from which the primary care data are obtained, are required, share relevant health information, support the public health response, the pandemic, and have been informed of the OpenSAFELY analytics platform.

https://github.com/opensafely/immunosuppressant-meds-research

## ADMINISTRATIVE

### Acknowledgements

We are very grateful for all the support received from the TPP Technical Operations team throughout this work, and for generous assistance from the information governance and database teams at NHS England / NHSX. Additionally the North East Commissioning Support Unit provided support on behalf of all Commissioning Support Unit to aggregate the high cost drugs data. This study uses electronic health records, data are provided by patients and collected by the National Health Service as part of their care and support. We are very grateful to Professor Joe West from the University of Nottingham and Professor Daniel Prieto-Alhmabra for comments on an early version of the protocol.

### Funding

TPP and NECS CSU provided technical expertise and data infrastructure centre *pro bono* in the context of a national emergency. This work was supported by the Medical Research Council MR/V015737/1. BG’s work on better use of data in healthcare more broadly is currently funded in part by: NIHR Oxford Biomedical Research Centre, NIHR Applied Research Collaboration Oxford and Thames Valley, the Mohn-Westlake Foundation, NHS England, and the Health Foundation; all DataLab staff are supported by BG’s grants on this work. LS reports grants from Wellcome, MRC, NIHR, UKRI, British Council, GSK, British Heart Foundation, and Diabetes UK outside this work. JB is funded by a studentship from GSK. AS is employed by LSHTM on a fellowship sponsored by GSK. KB holds a Sir Henry Dale fellowship jointly funded by Wellcome and the Royal Society. HIM is funded by the National Institute for Health Research (NIHR) Health Protection Research Unit in Immunisation, a partnership between Public Health England and LSHTM. AYSW holds a fellowship from BHF. EW holds grants from MRC. ID holds grants from NIHR and GSK. RM holds a Sir Henry Wellcome Fellowship funded by the Wellcome Trust. HF holds a UKRI fellowship. RME is funded by HDR-UK and the MRC. SML was supported by a Wellcome Trust Senior Research Fellowship in Clinical Science (205039/Z/16/Z). The findings and conclusions in this report are those of the authors and do not necessarily represent the views of the funders. SML was also supported by Health Data Research UK (Grant number: LOND1), which is funded by the UK Medical Research Council, Engineering and Physical Sciences Research Council, Economic and Social Research Council, Department of Health and Social Care (England), Chief Scientist Office of the Scottish Government Health and Social Care Directorates, Health and Social Care Research and Development Division (Welsh Government), Public Health Agency (Northern Ireland), British Heart Foundation and Wellcome Trust. SML is an investigator on the European Union Horizon 2020-funded BIOMAP Consortium (http://www.biomap-imi.eu/). CS acknowledges support for this research from the NIHR Biomedical Research Centre (BRC) at King’s College London and Guy’s and St Thomas’ NHS Foundation Trust and the Psoriasis Association. CWL is funded by a UKRI Future Leaders Fellowship.

The views expressed are those of the authors and not necessarily those of the NIHR, NHS England, Public Health England or the Department of Health and Social Care.

Funders had no role in the study design, collection, analysis, and interpretation of data; in the writing of the report; and in the decision, submit the article for publication.

This research was funded in whole or in part by the Wellcome Trust [G205039/Z/16/Z]. For the purpose of Open Access, the author has applied a CC BY public copyright licence to any Author Accepted Manuscript (AAM) version arising from this submission

### Information governance and ethical approval

NHS England is the data controller; *TPP is the data processor*; and the key researchers on OpenSAFELY are acting on behalf of NHS England. This implementation of OpenSAFELY is hosted within the *TPP environment which is* accredited, the ISO 27001 information security standard and *is* NHS IG Toolkit complian^t;31,32^ patient data has been pseudonymised for analysis and linkage using industry standard cryptographic hashing techniques; all pseudonymised datasets transmitted for linkage onto OpenSAFELY are encrypted; access, the platform is via a virtual private network (VPN) connection, restricted, a small group of researchers; the researchers hold contracts with NHS England and only access the platform, initiate database queries and statistical models; all database activity is logged; only aggregate statistical outputs leave the platform environment following best practice for anonymisation of results such as statistical disclosure control for low cell counts.^33^ The OpenSAFELY research platform adheres, the obligations of the UK General Data Protection Regulation (GDPR) and the Data Protection Act 2018. In March 2020, the Secretary of State for Health and Social Care used powers under the UK Health Service (Control of Patient Information) Regulations 2002 (COPI), require organisations, process confidential patient information for the purposes of protecting public health, providing healthcare services, the public and monitoring and managing the COVID-19 outbreak and incidents of exposure; this sets aside the requirement for patient consen^t.34^ Taken together, these provide the legal bases, link patient datasets on the OpenSAFELY platform. GP practices, from which the primary care data are obtained, are required, share relevant health information, support the public health response, the pandemic, and have been informed of the OpenSAFELY analytics platform.

This study was approved by the Health Research Authority (REC Reference 20/LO/0651) and by the LSHTM Ethics Board (Reference 21863).

### Guarantor

BG/LS is guarantor.

### Contributions

BG conceived the OpenSAFELY platform and the approach.

LS and BG led the project overall and are guarantors.

SML, CS, NAK, CL, JG, KEM, BMK, SJWE conceptualised the study.

SB led on software development.

AM led on information governance.

CJB, CEM, DH, RC, GH, TW, SCJB, PI, JC, DE, JP, and SH curated data;

SML, CS, NAK, CL, JG, KEM, BMK, KB, CTR, BM, CJB, CEM, IJD, AJW, HIM, JC, HJF, HJC, JT, RME, LAT, RME, AYSW and JP conceptualised disease categories and code lists;

MY, SN, JG, KSB, JB and JM wrote statistical analysis code;

KEM, KSB, JG, SN, MY, JB did data visualisation;

EJW, HJC, LS, and BG obtained ethical approvals;

CJB, CEM, SCJB, SD, AG, LF, PI, AJW, JC, DE, WJH, and FH contributed to software development;

and AS, SML, KEM, DH, CS, KSB, JG, SN, MY, NAK, JM, LAT, RME, AYSW and SJWE reviewed and edited the manuscript.

All authors were involved in design and conceptual development and reviewed and approved the final manuscript.

## SUPPLEMENTARY MATERIAL

### Text S1. High-cost drugs dataset

To achieve a comprehensive national medicines dataset, we arranged for the unique collation of a single national hospital medication dataset for “high-cost drugs”. High-cost drugs are typically newer and more expensive medications (e.g., adalimumab) that are supplied by hospitals in England for ongoing disease management. High-cost drugs are paid for by commissioners (in England, National Health Service Clinical Commission Groups commission most secondary care services and play a part in the commissioning of GP services) and as part of this process, unique patient identifiers are supplied by hospitals, the responsible commissioner (there are approximately 200 commissioners). The North East Commissioning Support Unit collected high-cost drug data from commissioners and compiled it into a single national dataset (including medication name, date of medication supply and a unique patient identifier – NHS number – allowing linkage with other healthcare data). To our knowledge this is the first time in England that a comprehensive national medicines dataset including high-cost drug data has been available at individual patient level. More information is available in our accompanying short data report.[link will be provided in published paper]

Drug exposure was defined by at least one prescription, or delivery of medication to an individual before March 1^st^, 2020 (date chosen as some of medications were either specifically used, or stopped, due to the pandemic). For each individual, we defined drug exposure based on the closest drug recorded prior to the study start (March 1^st^ 2020) allowing for a maximum of 6 months before the start of the study for all agents apart from rituximab, where we permitted a 12-month exposure window (to allow for the frequency of treatment and the longer duration of response of rituximab, including known prolonged effects on vaccine responses).^16,17^ We could not evaluate medication switching or adherence during the follow-up period, as data was only available up to March 2020.

### Text S2. Covariates

All analytical code for all covariates definitions are openly available online for inspection and re-use. Some brief descriptions are provided below.

#### Age

Age was defined by the age reached by the 1^st^ March 2020 (study start date). We categorised age groups as: 18–39, 40–49, 50–59, 60–69, 70–79 and 80+ years.

#### Smoking and body mass index

We categorised smoking status as current-, former- or never-smokers, based on primary care morbidity coding recorded prior to March 1^st^, 2020. People with missing smoking status were categorised as never smokers.

Body mass index (BMI) was calculated from height and weight measures recorded in primary care, based on weight measurements within the last 10 years (before March 1^st^, 2020), and restricted to records from after age 16 years. BMI was categorised according to World Health Organisation (WHO) BMI classification: no evidence of obesity (BMI < 30); obese class I (BMI 30–34.9); obese class II (BMI 35–39.9); and obese class III (BMI 40+). Individuals with missing BMI data were categorised as having no evidence of obesity (i.e., BMI < 30).

We undertook a sensitivity analysis without re-categorising missing BMI and smoking status data (i.e., complete-case analysis of only those with complete smoking and BMI data). Our approach reflects an awareness that BMI and smoking data are likely to be not missing at random, and has precedent in previous OpenSAFELY analyses.^1,2^

#### Current glucocorticoid use

We defined current glucocorticoid use based on a primary care prescription of oral prednisolone at any dose in the three months before study start.

#### Diabetes mellitus

We defined diabetes mellitus based on the most recent glycosylated haemoglobin (Hba1c) measurement recorded in primary care at any time before study start (March 1^st^, 2020): controlled diabetes Hba1c<58mmol/mol; uncontrolled diabetes Hba1c≥58mmol/mol; or diabetes (based on morbidity coded diabetes) with no recorded Hba1c measure.

#### End stage renal failure

We defined end stage renal failure based on estimated glomerular filtration rate calculated from the most recent serum creatinine result recorded 15 months before study start (March 1^st^, 2020) in primary care data. We only adjusted for end stage renal failure in sensitivity analyses.

#### Ethnicity

We defined ethnicity based on coding in primary care. We classified ethnicity as: White, Black, South Asian, and mixed or other. We only adjusted for ethnicity in sensitivity analyses due to the large proportion of missing data (**Tables 1 and 2**).

#### Other chronic comorbidities

We defined cardiovascular disease (chronic heart failure, ischaemic heart disease, and severe valve or congenital heart disease likely to require lifelong follow-up), cancer (excluding non-melanoma skin cancer), and stroke based on any morbidity coding in primary care recorded prior to March 1^st^, 2020 (study start date). We also adjusted for chronic respiratory disease (chronic obstructive pulmonary disease, fibrosing lung disease, bronchiectasis or cystic fibrosis) and chronic liver disease in a sensitivity analysis, again defined based on any morbidity coding in primary care recorded prior to study start.

**References**

1. Bhaskaran K, Bacon S, Evans SJ, et al. Factors associated with deaths due to COVID-19 versus other causes: population-based cohort analysis of UK primary care data and linked national death registrations within the OpenSAFELY platform. Lancet Reg Health Eur 2021;6:100109.

2. Bhaskaran K, Rentsch CT, MacKenna B, et al. HIV infection and COVID-19 death: a population-based cohort analysis of UK primary care data and linked national death registrations within the OpenSAFELY platform. Lancet HIV 2021;8:e24-e32

**Figure S1.**
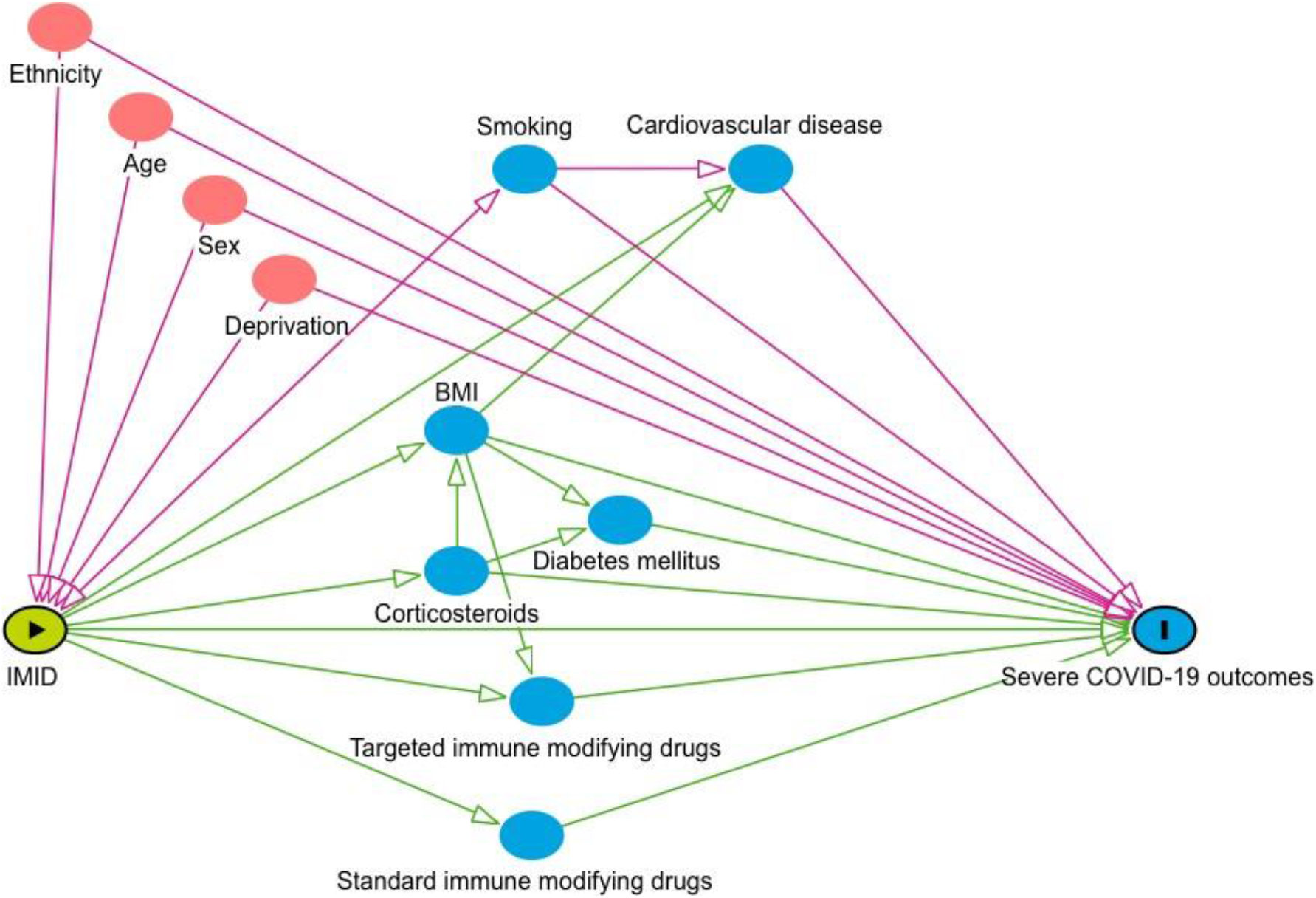
Conceptual framework: risk of COVID-19-related death in people with IMIDs compared to the general population. The conceptual framework represents the assumed associations between covariates and primary exposure and outcome. Pink circles represent ancestors of the exposure and outcome, blue circles represent ancestors of the outcome, pink lines represent biasing paths (i.e. confounding) and green lines represent causal paths.. The minimally sufficient adjustment set (i.e. the covariates adjusted for in confounder adjusted models) represents covariates such that the adjustment for this set of variables will minimize confounding bias when estimating the association between the exposure and the outcome.

**Figure S2.**
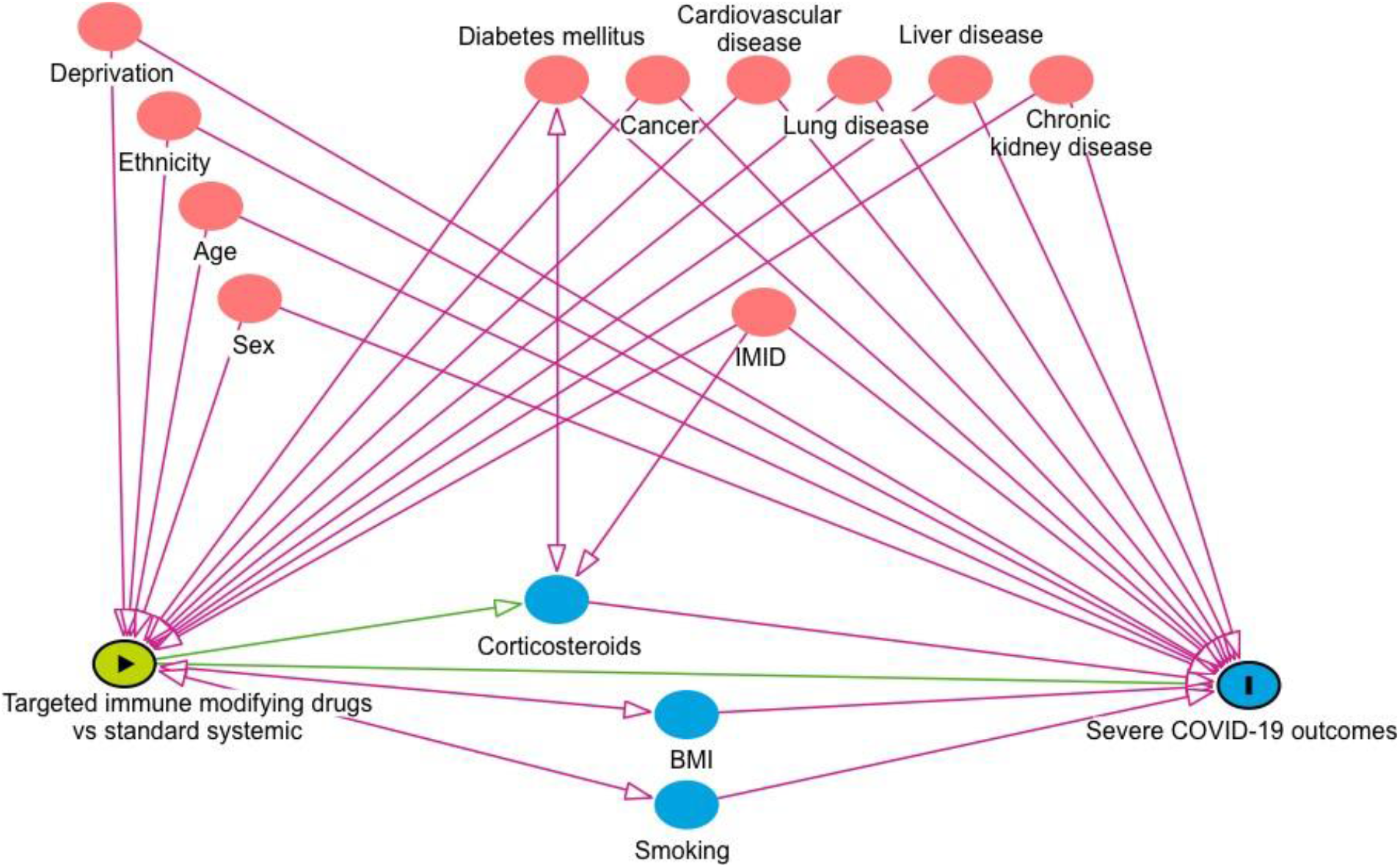
Conceptual framework: risk of COVID-19-related death in people with IMIDS taking targeted immunosuppressive drugs compared to those prescribed standard systemic drugs. The conceptual framework A directed acyclic graph represents the assumed associations between covariates and primary exposure and outcome. Pink circles represent ancestors of the exposure and outcome, blue circles representancestors of the outcome, pink lines represent biasing paths (i.e. confounding) and green lines represent causal paths. The minimally sufficient adjustment set (i.e. the covariates adjusted for in confounder adjusted models) represents covariates such that the adjustment for this set of variables will minimize confounding bias when estimating the association between the exposure and the outcome.

**Table S1.**
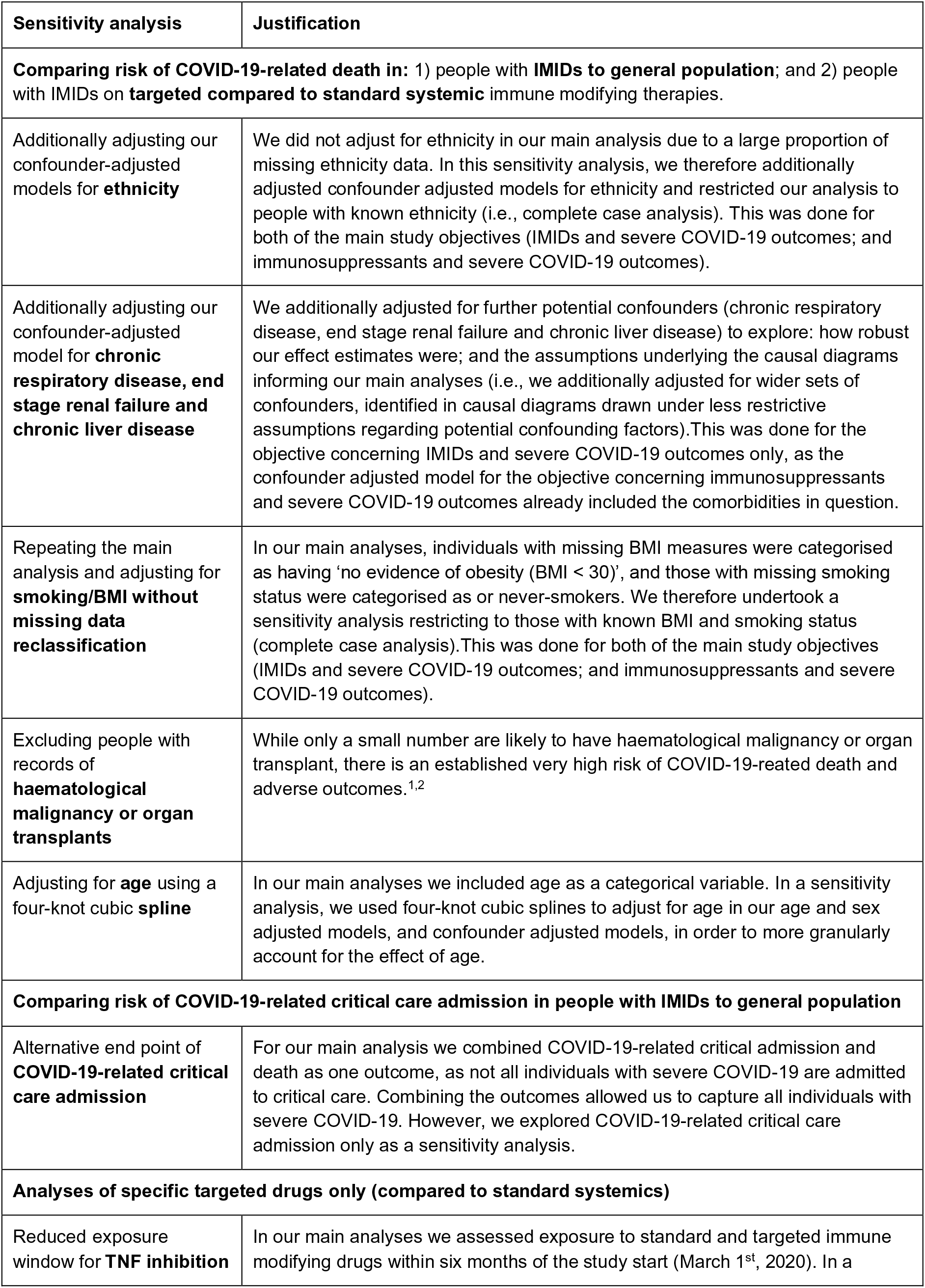
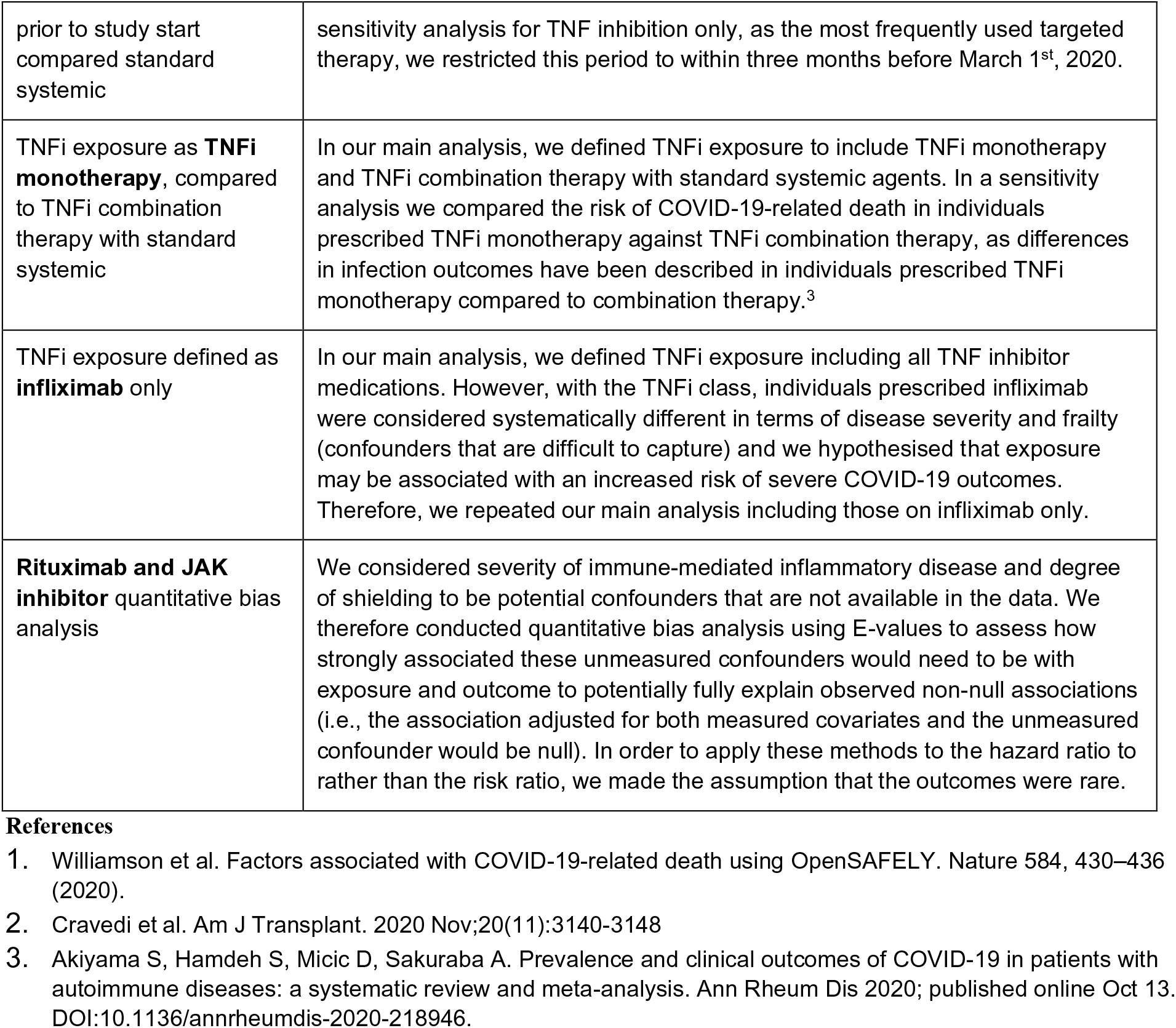
Sensitivity analyses.

| Sensitivity analysis | Justification |
| --- | --- |
| **Comparing risk of COVID-19-related death in: 1) people with IMIDs to general population; and 2) people with IMIDs on targeted compared to standard systemic immune modifying therapies.** |  |
| Additionally adjusting our confounder-adjusted models for **ethnicity** | We did not adjust for ethnicity in our main analysis due to a large proportion of missing ethnicity data. In this sensitivity analysis, we therefore additionally adjusted confounder adjusted models for ethnicity and restricted our analysis to people with known ethnicity (i.e., complete case analysis). This was done for both of the main study objectives (IMIDs and severe COVID-19 outcomes; and immunosuppressants and severe COVID-19 outcomes). |
| Additionally adjusting our confounder-adjusted model for **chronic respiratory disease, end stage renal failure and chronic liver disease** | We additionally adjusted for further potential confounders (chronic respiratory disease, end stage renal failure and chronic liver disease) to explore: how robust our effect estimates were; and the assumptions underlying the causal diagrams informing our main analyses (i.e., we additionally adjusted for wider sets of confounders, identified in causal diagrams drawn under less restrictive assumptions regarding potential confounding factors). This was done for the objective concerning IMIDs and severe COVID-19 outcomes only, as the confounder adjusted model for the objective concerning immunosuppressants and severe COVID-19 outcomes already included the comorbidities in question. |
| Repeating the main analysis and adjusting for **smoking/BMI without missing data reclassification** | In our main analyses, individuals with missing BMI measures were categorised as having 'no evidence of obesity (BMI < 30)', and those with missing smoking status were categorised as or never-smokers. We therefore undertook a sensitivity analysis restricting to those with known BMI and smoking status (complete case analysis). This was done for both of the main study objectives (IMIDs and severe COVID-19 outcomes; and immunosuppressants and severe COVID-19 outcomes). |
| Excluding people with records of **haematological malignancy or organ transplants** | While only a small number are likely to have haematological malignancy or organ transplant, there is an established very high risk of COVID-19-related death and adverse outcomes.<sup>1,2</sup> |
| Adjusting for **age** using a four-knot cubic **spline** | In our main analyses we included age as a categorical variable. In a sensitivity analysis, we used four-knot cubic splines to adjust for age in our age and sex adjusted models, and confounder adjusted models, in order to more granularly account for the effect of age. |
| **Comparing risk of COVID-19-related critical care admission in people with IMIDs to general population** |  |
| Alternative end point of **COVID-19-related critical care admission** | For our main analysis we combined COVID-19-related critical admission and death as one outcome, as not all individuals with severe COVID-19 are admitted to critical care. Combining the outcomes allowed us to capture all individuals with severe COVID-19. However, we explored COVID-19-related critical care admission only as a sensitivity analysis. |
| **Analyses of specific targeted drugs only (compared to standard systemics)** |  |
| Reduced exposure window for **TNF inhibition** | In our main analyses we assessed exposure to standard and targeted immune modifying drugs within six months of the study start (March 1<sup>st</sup>, 2020). In a |
| prior to study start compared standard systemic | sensitivity analysis for TNF inhibition only, as the most frequently used targeted therapy, we restricted this period to within three months before March 1<sup>st</sup>, 2020. |
| TNFi exposure as **TNFi monotherapy**, compared to TNFi combination therapy with standard systemic | In our main analysis, we defined TNFi exposure to include TNFi monotherapy and TNFi combination therapy with standard systemic agents. In a sensitivity analysis we compared the risk of COVID-19-related death in individuals prescribed TNFi monotherapy against TNFi combination therapy, as differences in infection outcomes have been described in individuals prescribed TNFi monotherapy compared to combination therapy.<sup>3</sup> |
| TNFi exposure defined as **infliximab** only | In our main analysis, we defined TNFi exposure including all TNF inhibitor medications. However, with the TNFi class, individuals prescribed infliximab were considered systematically different in terms of disease severity and frailty (confounders that are difficult to capture) and we hypothesised that exposure may be associated with an increased risk of severe COVID-19 outcomes. Therefore, we repeated our main analysis including those on infliximab only. |
| **Rituximab and JAK inhibitor** quantitative bias analysis | We considered severity of immune-mediated inflammatory disease and degree of shielding to be potential confounders that are not available in the data. We therefore conducted quantitative bias analysis using E-values to assess how strongly associated these unmeasured confounders would need to be with exposure and outcome to potentially fully explain observed non-null associations (i.e., the association adjusted for both measured covariates and the unmeasured confounder would be null). In order to apply these methods to the hazard ratio to rather than the risk ratio, we made the assumption that the outcomes were rare. |

**Figure S3.**
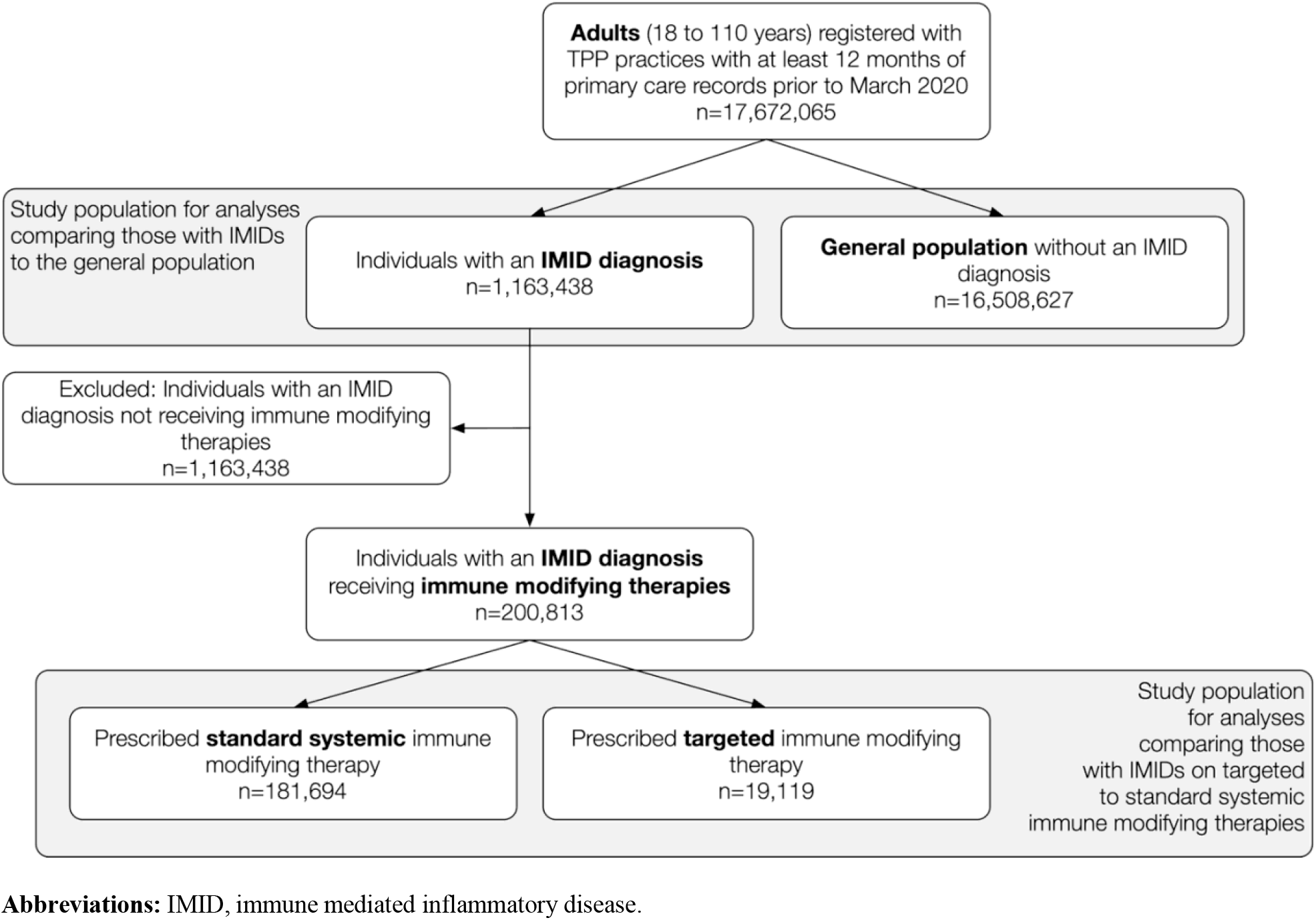
Flow diagram for identification of study population.

**Table S2.** **Main analysis:** Hazard Ratios (HRs) and 95% confidence intervals (CI) for COVID-19-related death, death/critical care admission or hospitalisation, in people with **IMIDs compared to the general population**

|  | Events | Person-years | Rate per 1000 PY (95%CI) | Crude HR (95%CI) | Age and sex adjusted HR (95%CI) | Confounder adjusted HR (95%CI) | Mediator adjusted HR (95% CI) |
| --- | --- | --- | --- | --- | --- | --- | --- |
| **COVID-19 death** |  |  |  |  |  |  |  |
| General population | 40,453 | 8,293,798 | 4.88 (4.83, 4.93) | REF | REF | REF | REF |
| All IMID | 4,824 | 583,617 | 8.27 (8.03, 8.50) | 1.70 (1.65, 1.75) | 1.27 (1.23, 1.31) | 1.23 (1.20, 1.27) | 1.15 (1.11, 1.18) |
| Inflammatory joint disease | 1,856 | 136,333 | 13.61 (13.00, 14.25) | 2.75 (2.62, 2.88) | 1.51 (1.44, 1.58) | 1.47 (1.40, 1.54) | 1.30 (1.24, 1.37) |
| Inflammatory skin disease | 2,608 | 386,455 | 6.75 (6.49, 7.01) | 1.34 (1.29, 1.40) | 1.16 (1.11, 1.20) | 1.12 (1.08, 1.17) | 1.07 (1.02, 1.11) |
| Inflammatory bowel disease | 721 | 99,875 | 7.22 (6.70, 7.77) | 1.42 (1.32, 1.53) | 1.15 (1.07, 1.24) | 1.12 (1.04, 1.21) | 1.07 (0.99, 1.15) |
| **COVID-19 critical care admission or death** |  |  |  |  |  |  |  |
| General population | 43,972 | 8,311,564 | 5.29 (5.24, 5.34) | REF | REF | REF | REF |
| All IMID | 5,208 | 585,538 | 8.89 (8.65, 9.14) | 1.68 (1.63, 1.73) | 1.28 (1.24, 1.31) | 1.24 (1.21, 1.28) | 1.16 (1.12, 1.19) |
| Inflammatory joint disease | 1,950 | 137,026 | 14.23 (13.61, 14.88) | 2.64 (2.52, 2.76) | 1.50 (1.43, 1.57) | 1.46 (1.39, 1.52) | 1.30 (1.24, 1.36) |
| Inflammatory skin disease | 2,867 | 387,508 | 7.40 (7.13, 7.67) | 1.36 (1.31, 1.41) | 1.18 (1.13, 1.22) | 1.15 (1.10, 1.19) | 1.08 (1.04, 1.13) |
| Inflammatory bowel disease | 784 | 100,186 | 7.83 (7.29, 8.39) | 1.42 (1.33, 1.53) | 1.16 (1.08, 1.24) | 1.13 (1.06, 1.22) | 1.08 (1.01, 1.16) |
| **COVID-19 hospitalisation** |  |  |  |  |  |  |  |
| General population | 72,862 | 8,309,954 | 8.77 (8.70, 8.83) | REF | REF | REF | REF |
| All IMID | 8,376 | 585,324 | 14.31 (14.01, 14.62) | 1.63 (1.60, 1.67) | 1.34 (1.31, 1.37) | 1.32 (1.29, 1.35) | 1.20 (1.17, 1.23) |
| Inflammatory joint disease | 2,869 | 136,962 | 20.95 (20.19, 21.62) | 2.34 (2.26, 2.43) | 1.57 (1.51, 1.63) | 1.53 (1.47, 1.59) | 1.32 (1.27, 1.37) |
| Inflammatory skin disease | 4,752 | 387,377 | 12.27 (11.92, 12.62) | 1.36 (1.33, 1.41) | 1.22 (1.19, 1.26) | 1.21 (1.17, 1.24) | 1.10 (1.07, 1.14) |
| Inflammatory bowel disease | 1,426 | 100,146 | 14.24 (13.51, 15.00) | 1.57 (1.49, 1.65) | 1.35 (1.28, 1.42) | 1.31 (1.24, 1.38) | 1.25 (1.19, 1.31) |

Planned comparisons were made between people with IMIDs, and IMID types (joint, bowel, skin), using the general population as the reference group.
**Confounder adjusted:** age, sex, deprivation, and smoking status
**Mediator adjusted:** age, sex, deprivation, smoking status, BMI, cardiovascular disease, diabetes mellitus and current glucocorticoid use
**Abbreviations:** HR, hazard ratio; CI, confidence interval; PY, person years; IMID, immune mediated inflammatory disease; BMI, body mass index.

**Table S3.**
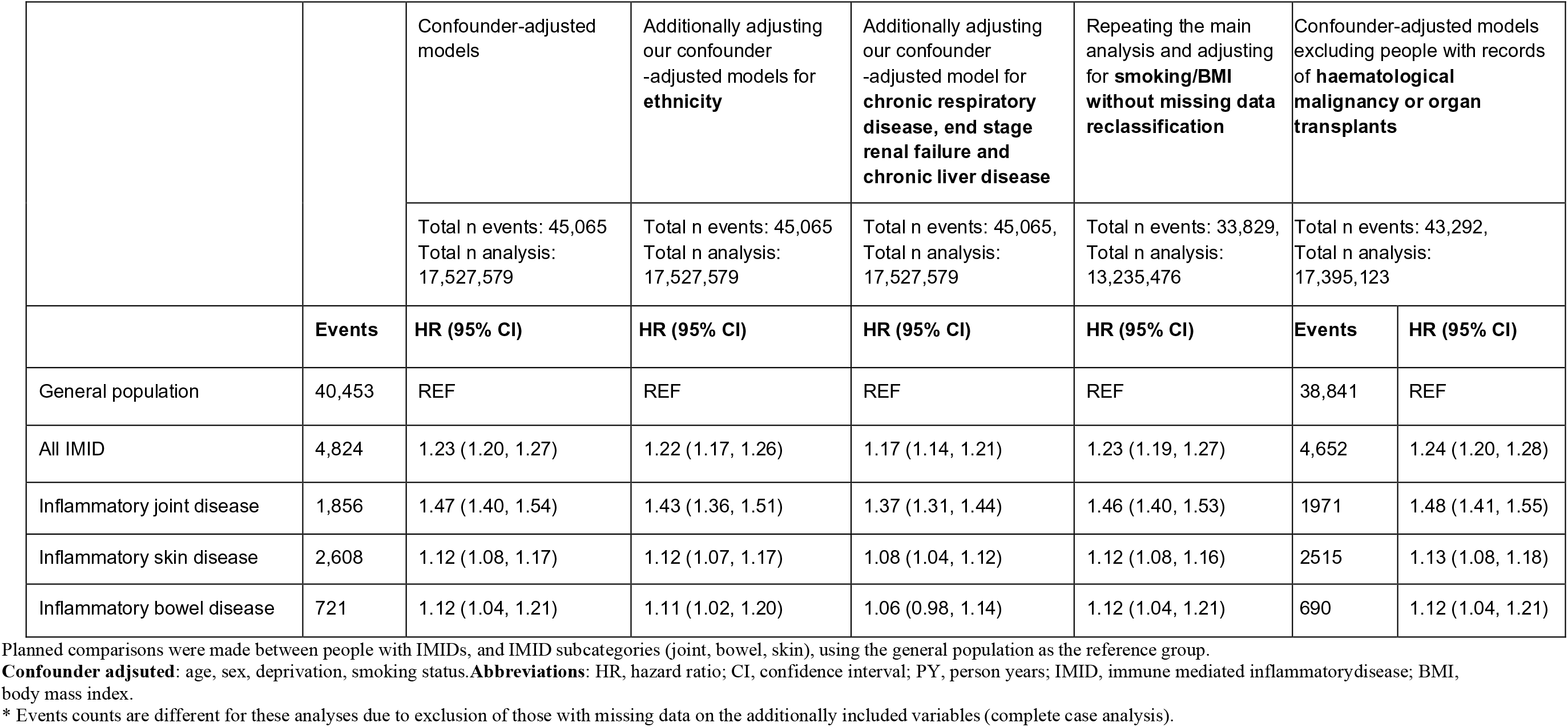
**Sensitivity analyses**: **COVID-19-related death** in **IMIDs** compared to the general population.

|  |  | Confounder-adjusted models | Additionally adjusting our confounder -adjusted models for **ethnicity** | Additionally adjusting our confounder -adjusted model for **chronic respiratory disease, end stage renal failure and chronic liver disease** | Repeating the main analysis and adjusting for **smoking/BMI without missing data reclassification** | Confounder-adjusted models excluding people with records of **haematological malignancy or organ transplants** |  |
| --- | --- | --- | --- | --- | --- | --- | --- |
|  |  | Total n events: 45,065<br>Total n analysis: 17,527,579 | Total n events: 45,065<br>Total n analysis: 17,527,579 | Total n events: 45,065,<br>Total n analysis: 17,527,579 | Total n events: 33,829,<br>Total n analysis: 13,235,476 | Total n events: 43,292,<br>Total n analysis: 17,395,123 |  |
|  | Events | HR (95% CI) | HR (95% CI) | HR (95% CI) | HR (95% CI) | Events | HR (95% CI) |
| General population | 40,453 | REF | REF | REF | REF | 38,841 | REF |
| All IMID | 4,824 | 1.23 (1.20, 1.27) | 1.22 (1.17, 1.26) | 1.17 (1.14, 1.21) | 1.23 (1.19, 1.27) | 4,652 | 1.24 (1.20, 1.28) |
| Inflammatory joint disease | 1,856 | 1.47 (1.40, 1.54) | 1.43 (1.36, 1.51) | 1.37 (1.31, 1.44) | 1.46 (1.40, 1.53) | 1971 | 1.48 (1.41, 1.55) |
| Inflammatory skin disease | 2,608 | 1.12 (1.08, 1.17) | 1.12 (1.07, 1.17) | 1.08 (1.04, 1.12) | 1.12 (1.08, 1.16) | 2515 | 1.13 (1.08, 1.18) |
| Inflammatory bowel disease | 721 | 1.12 (1.04, 1.21) | 1.11 (1.02, 1.20) | 1.06 (0.98, 1.14) | 1.12 (1.04, 1.21) | 690 | 1.12 (1.04, 1.21) |

Planned comparisons were made between people with IMIDs, and IMID subcategories (joint, bowel, skin), using the general population as the reference group.
**Confounder adjusted:** age, sex, deprivation, smoking status. **Abbreviations:** HR, hazard ratio; CI, confidence interval; PY, person years; IMID, immune mediated inflammatory disease; BMI, body mass index.
\* Events counts are different for these analyses due to exclusion of those with missing data on the additionally included variables (complete case analysis).

**Table S4.** **Sensitivity analyses**: **COVID-19-related critical care admission/death** in **IMIDs** compared tothe general population.

|  | COVID-19 death or critical care admission<br>Confounder adjusted analysis |  | COVID-19 critical care admission alternative outcome<br>Confounder adjusted analysis |  |
| --- | --- | --- | --- | --- |
|  | Events | Confounder Adjusted HR (95%CI) | Events | Confounder adjusted HR (95%CI) |
| General population | 43,972 | REF | 6070 | REF |
| All IMID | 5,208 | 1.24 (1.21, 1.28) | 679 | 1.36 (1.25, 1.47) |
| Inflammatory joint disease | 1,950 | 1.46 (1.39, 1.52) | 213 | 1.59 (1.38, 1.82) |
| Inflammatory skin disease | 2,867 | 1.15 (1.10, 1.19) | 431 | 1.33 (1.21, 1.47) |
| Inflammatory bowel disease | 784 | 1.13 (1.06, 1.22) | 96 | 1.05 (0.86, 1.29) |

Planned comparisons were made between people with IMIDs, and IMID type (joint, bowel, skin), using the general population as the reference group.
**Confounder adjusted:** age, sex, deprivation, and smoking status
**Abbreviations:** HR, hazard ratio; CI, confidence interval; PY, person years; IMID, immune mediated inflammatory disease; BMI, body mass index.

**Table S5.**
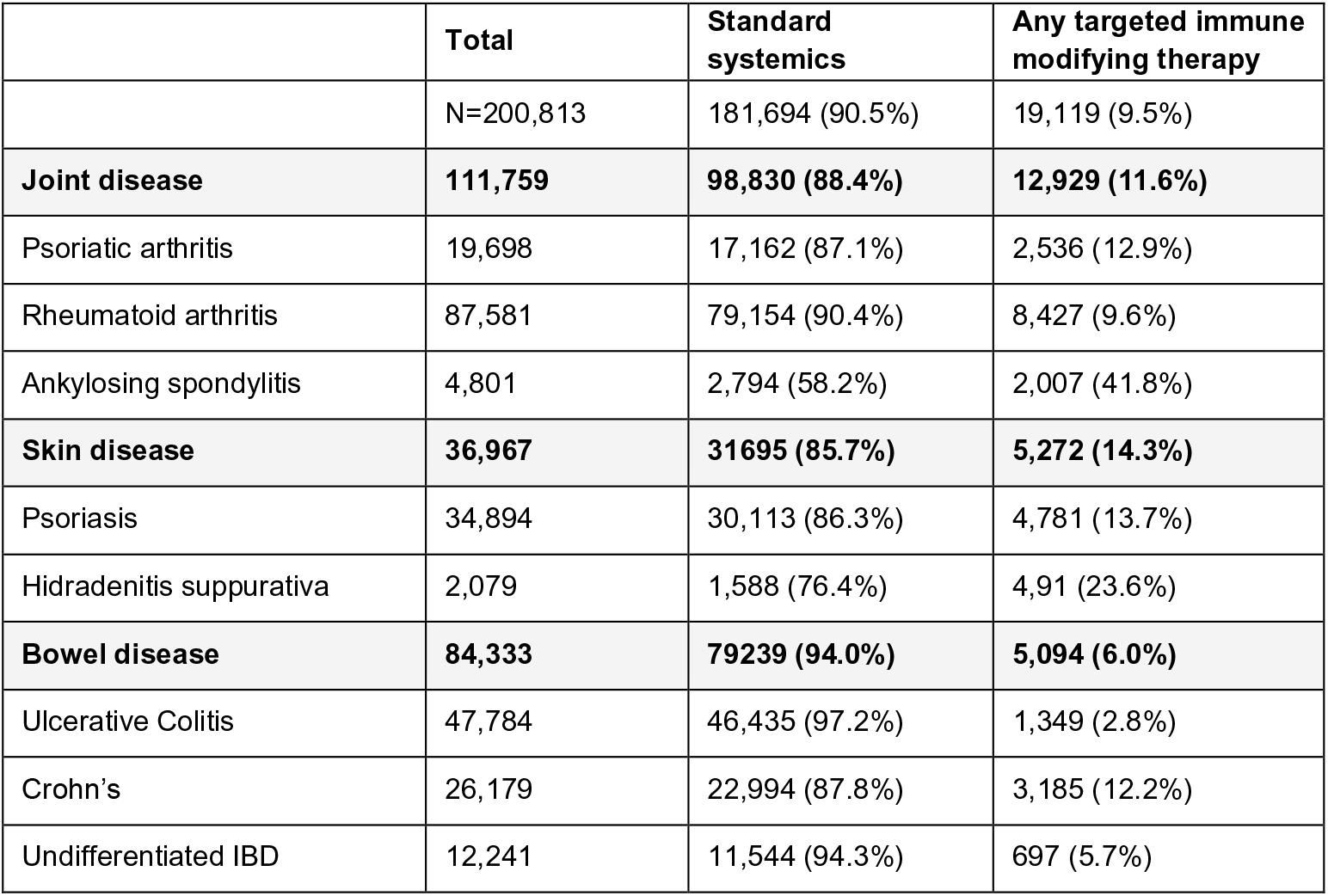
Number of individuals with each IMID type receiving standard systemic or any targeted immune modifying therapy

|  | **Total** | **Standard systemics** | **Any targeted immune modifying therapy** |
| --- | --- | --- | --- |
|  | N=200,813 | 181,694 (90.5%) | 19,119 (9.5%) |
| **Joint disease** | **111,759** | **98,830 (88.4%)** | **12,929 (11.6%)** |
| Psoriatic arthritis | 19,698 | 17,162 (87.1%) | 2,536 (12.9%) |
| Rheumatoid arthritis | 87,581 | 79,154 (90.4%) | 8,427 (9.6%) |
| Ankylosing spondylitis | 4,801 | 2,794 (58.2%) | 2,007 (41.8%) |
| **Skin disease** | **36,967** | **31695 (85.7%)** | **5,272 (14.3%)** |
| Psoriasis | 34,894 | 30,113 (86.3%) | 4,781 (13.7%) |
| Hidradenitis suppurativa | 2,079 | 1,588 (76.4%) | 4,91 (23.6%) |
| **Bowel disease** | **84,333** | **79239 (94.0%)** | **5,094 (6.0%)** |
| Ulcerative Colitis | 47,784 | 46,435 (97.2%) | 1,349 (2.8%) |
| Crohn's | 26,179 | 22,994 (87.8%) | 3,185 (12.2%) |
| Undifferentiated IBD | 12,241 | 11,544 (94.3%) | 697 (5.7%) |

**Table S6.**
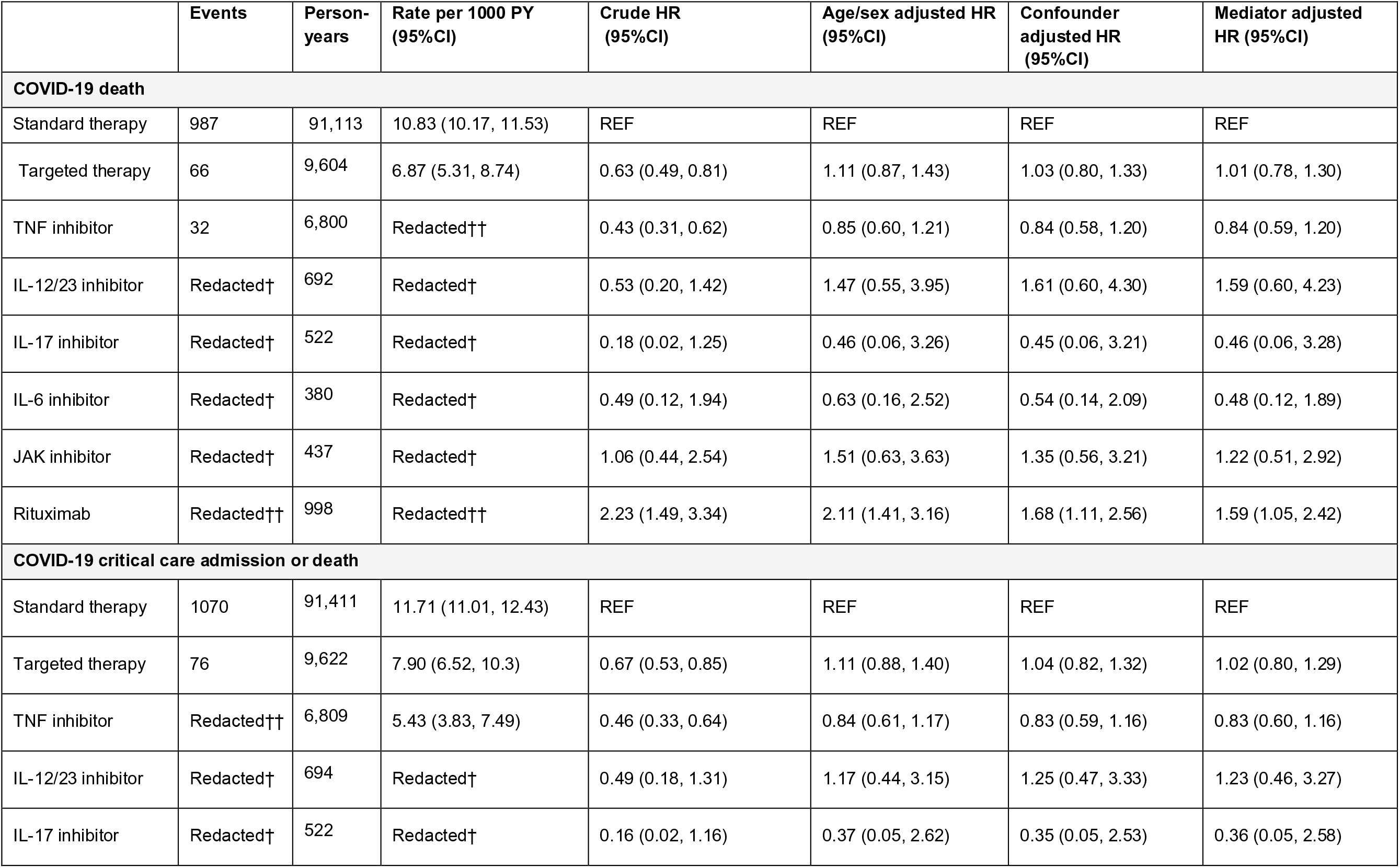
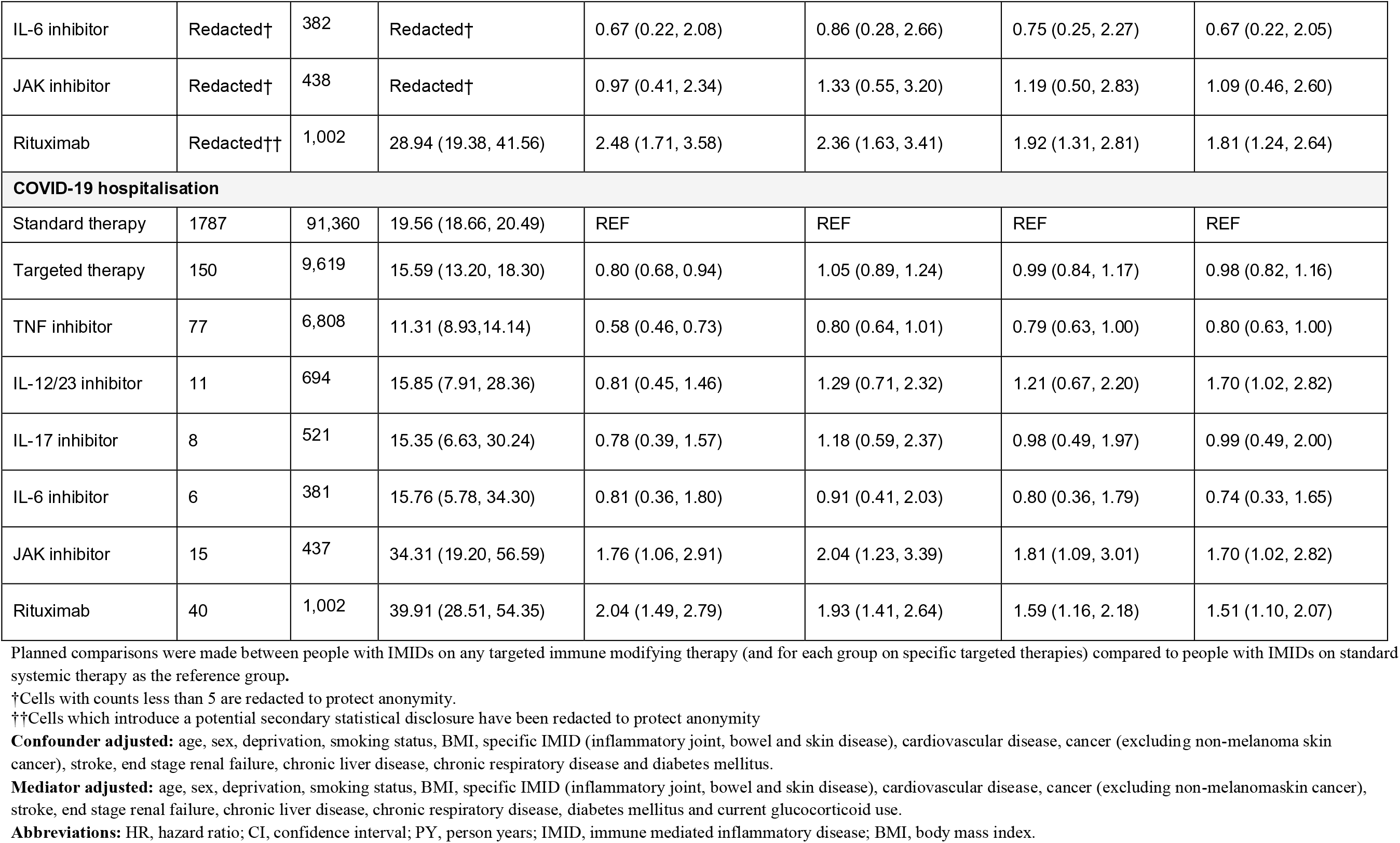
**Main analysis:** Hazard Ratios (HRs) and 95% confidence intervals (CI) for COVID-19-related death, death/critical care admission or hospitalisation, in people with **IMIDs on targeted immune modifying therapies** compared to those with **IMIDs on standard systemic immune modifying therapies**.

**Table S7.**
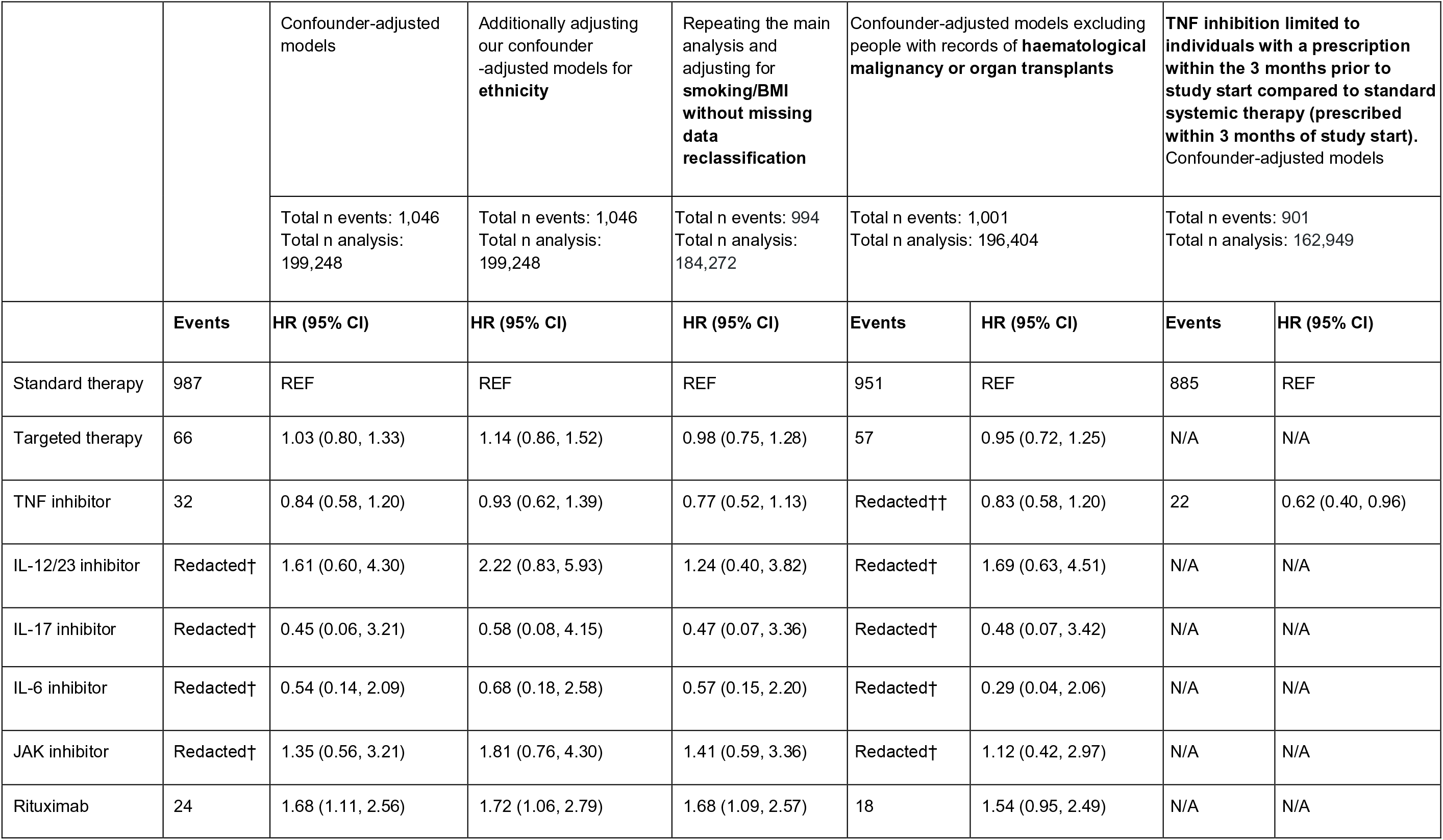
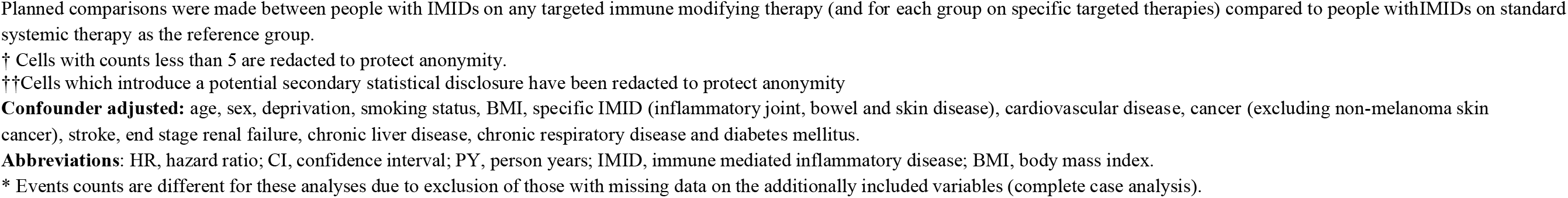
**Sensitivity analyses:** for **COVID-19-related death** in people with IMIDs on **targeted immune modifying therapies** compared tothose with IMIDs on standard systemic immune modifying therapies.

**Table S8.** **Sensitivity analyses**: **COVID-19-related critical care admission/death** in people with IMIDs on **targeted immune modifying therapies** compared to those with IMIDs on standard systemicimmune modifying therapies

|  | COVID-19 death or critical care admission<br>Confounder adjusted |  | COVID-19 critical care admission<br>alternative outcome<br>Confounder adjusted |  |
| --- | --- | --- | --- | --- |
|  | Events | Adjusted HR<br>(95%CI) | Events | Adjusted HR<br>(95%CI) |
| Standard therapy | 1070 | REF | 172 | REF |
| Targeted therapy | 76 | 1.04 (0.82, 1.32) | 17 | 0.90 (0.54, 1.49) |
| TNF inhibitor | Redacted†† | 0.83 (0.59, 1.16) | 8 | 0.63 (0.31, 1.30) |
| IL-12/23 inhibitor | Redacted† | 1.25 (0.47, 3.33) | Redacted† | 0.91 (0.12, 6.61) |
| IL-17 inhibitor | Redacted† | 0.35 (0.05, 2.53) | Redacted† | 0.83 (0.11, 6.03) |
| IL-6 inhibitor | Redacted† | 0.75 (0.25, 2.27) | Redacted† | 2.44 (0.60, 10.02) |
| JAK inhibitor | Redacted† | 1.19 (0.50, 2.83) | 0 | 0 |
| Rituximab | Redacted†† | 1.92 (1.31, 2.81) | 7 | 2.69 (1.24, 5.85) |

Planned comparisons were made between people with IMIDs on any targeted immune modifying therapy (and for each group on specific targeted therapies) compared to people with IMIDs on standard systemic therapy as the reference group.
† Cells with counts less than 5 are redacted to protect anonymity.
†† Cells which introduce a potential secondary statistical disclosure have been redacted to protect anonymity
**Abbreviations:** HR, hazard ratio; CI, confidence interval; PY, person years; IMID, immune mediated inflammatory disease; BMI, body mass index.

**Table S9.** **Sensitivity analysis:** Hazard Ratios (95% CI) for COVID-19-related death with **TNF inhibition monotherapy** compared to those receiving combination TNF inhibition and standard systemic therapy.

|  | Events | Confounder adjusted HR (95%CI) |
| --- | --- | --- |
| TNF and standard systemic combination therapy | 21 | REF |
| TNF inhibitor monotherapy | 11 | 0.58 (0.26, 1.34) |

**Abbreviations:** HR, hazard ratio; CI, confidence interval; PY, person years; IMID, immune mediated inflammatory disease; BMI, body mass index.

**Table S10.**
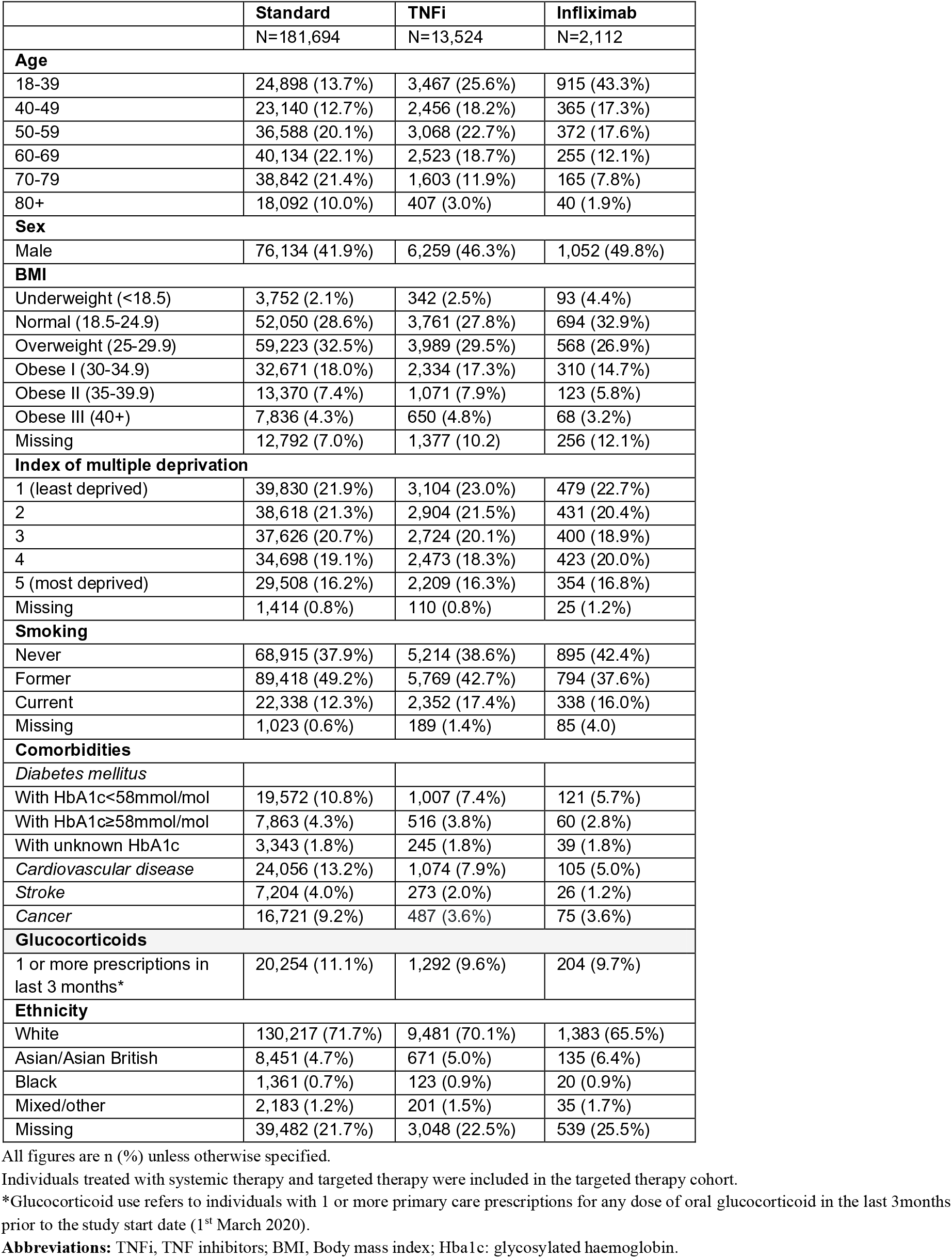
Descriptive characteristics of population on targeted immunosuppression with **infliximab**

**Table S11.** **Sensitivity analysis:** Hazard Ratios (95% CI) for **COVID-19-related death** in people with anIMID on **infliximab therapy** compared to those on standard systemic therapy

|  | Events | Confounder adjusted HR (95%CI) |
| --- | --- | --- |
| **Main analysis** |  |  |
| Standard therapy | 987 | REF |
| TNF inhibitor | 32 | 0.84 (0.58, 1.20) |
| **Sensitivity analysis with TNFi exposure defined as infliximab only** |  |  |
| Standard therapy | 987 | REF |
| Infliximab | Redacted # | 1.44 (0.60, 3.48) |

**Abbreviations:** HR, hazard ratio; CI, confidence interval.

**Table S12.** **Analysis adjusting for age using a four-knot cubic spline:** Hazard Ratios (HRs) and 95% confidence intervals (CI) for COVID-19-related death, death/critical care admission or hospitalisation, in people with **IMIDs compared to the general population**

|  | **Main analysis:**<br>**Age and sex**<br>**adjusted HR**<br>**(95%CI)** | **Main analysis:**<br>**Confounder**<br>**adjusted HR**<br>**(95%CI)** | **Age and sex**<br>**adjusted with**<br>**spline HR**<br>**(95%CI)** | **Confounder**<br>**adjusted HR with**<br>**spline**<br>**(95%CI)** |
| --- | --- | --- | --- | --- |
| **COVID-19 death** |  |  |  |  |
| General population | REF | REF | REF | REF |
| All IMID | 1.27 (1.23, 1.31) | 1.23 (1.20, 1.27) | 1.32 (1.28, 1.35) | 1.28 (1.24, 1.32) |
| Inflammatory joint disease | 1.51 (1.44, 1.58) | 1.47 (1.40, 1.54) | 1.58 (1.51, 1.66) | 1.54 (1.47, 1.61) |
| Inflammatory skin disease | 1.16 (1.11, 1.20) | 1.12 (1.08, 1.17) | 1.19 (1.14, 1.24) | 1.16 (1.11, 1.20) |
| Inflammatory bowel disease | 1.15 (1.07, 1.24) | 1.12 (1.04, 1.21) | 1.19 (1.10, 1.28) | 1.16 (1.08, 1.25) |
| **COVID-19 critical care admission or death** |  |  |  |  |
| General population | REF | REF | REF | REF |
| All IMID | 1.28 (1.24, 1.31) | 1.24 (1.21, 1.28) | 1.31 (1.28, 1.35) | 1.28 (1.24, 1.32) |
| Inflammatory joint disease | 1.50 (1.43, 1.57) | 1.46 (1.39, 1.52) | 1.56 (1.49, 1.63) | 1.51 (1.45, 1.58) |
| Inflammatory skin disease | 1.18 (1.13, 1.22) | 1.15 (1.10, 1.19) | 1.21 (1.16, 1.25) | 1.18 (1.13, 1.22) |
| Inflammatory bowel disease | 1.16 (1.08, 1.24) | 1.13 (1.06, 1.22) | 1.19 (1.11, 1.28) | 1.17 (1.09, 1.25) |
| **COVID-19 hospitalisation** |  |  |  |  |
| General population | REF | REF | REF | REF |
| All IMID | 1.34 (1.31, 1.37) | 1.32 (1.29, 1.35) | 1.34 (1.31, 1.37) | 1.33 (1.30, 1.36) |
| Inflammatory joint disease | 1.57 (1.51, 1.63) | 1.53 (1.47, 1.59) | 1.57 (1.51, 1.63) | 1.53 (1.48, 1.59) |
| Inflammatory skin disease | 1.22 (1.19, 1.26) | 1.21 (1.17, 1.24) | 1.22 (1.19, 1.26) | 1.21 (1.18, 1.25) |
| Inflammatory bowel disease | 1.35 (1.28, 1.42) | 1.31 (1.24, 1.38) | 1.35 (1.28, 1.42) | 1.31 (1.25, 1.38) |

Planned comparisons were made between people with IMIDs, and IMID types (joint, bowel, skin), using the general population as the reference group.
**Confounder adjusted:** age, sex, deprivation, and smoking status
**Abbreviations:** HR, hazard ratio; CI, confidence interval; PY, person years; IMID, immune mediated inflammatory disease; deprivation, index of multiple deprivation.

**Table S13.** **Analysis adjusting for age using a four-knot cubic spline:** Hazard Ratios (HRs) and 95% confidence intervals (CI) for COVID-19-related death, death/critical care admission or hospitalisation, in people with **IMIDs on targeted immune modifying therapies** compared tothose with **IMIDs on standard systemic immune modifying therapies**.

|  | Main analysis:<br>Age and sex<br>adjusted HR<br>(95%CI) | Main analysis:<br>Confounder<br>adjusted HR<br>(95%CI) | Using age spline:<br>Age and sex<br>adjusted HR<br>(95%CI) | Using age spline:<br>Confounder<br>adjusted HR<br>(95%CI) |
| --- | --- | --- | --- | --- |
| **COVID-19 death** |  |  |  |  |
| Standard therapy | REF | REF | REF | REF |
| Targeted therapy | 1.11 (0.87, 1.43) | 1.03 (0.80, 1.33) | 1.18 (0.92, 1.52) | 1.16 (0.90, 1.50) |
| TNF inhibitor | 0.85 (0.60, 1.21) | 0.84 (0.58, 1.20) | 0.49 (0.07, 3.48) | 0.90 (0.63, 1.29) |
| IL-12/23 inhibitor | 1.47 (0.55, 3.95) | 1.61 (0.60, 4.30) | 1.56 (0.58, 4.18) | 1.57 (0.59, 4.22) |
| IL-17 inhibitor | 0.46 (0.06, 3.26) | 0.45 (0.06, 3.21) | 0.68 (0.17, 2.72) | 0.50 (0.07, 3.57) |
| IL-6 inhibitor | 0.63 (0.16, 2.52) | 0.54 (0.14, 2.09) | 1.34 (0.55, 3.23) | 0.68 (0.17, 2.68) |
| JAK inhibitor | 1.51 (0.63, 3.63) | 1.35 (0.56, 3.21) | 1.54 (0.64, 3.70) | 1.56 (0.65, 3.74) |
| Rituximab | 2.11 (1.41, 3.16) | 1.68 (1.11, 2.56) | 2.24 (1.50, 3.36) | 2.16 (1.42, 3.26) |
| **COVID-19 critical care admission or death** |  |  |  |  |
| Standard therapy | REF | REF | REF | REF |
| Targeted therapy | 1.11 (0.88, 1.40) | 1.04 (0.82, 1.32) | 1.17 (0.93, 1.48) | 1.16 (0.91, 1.47) |
| TNF inhibitor | 0.84 (0.61, 1.17) | 0.83 (0.59, 1.16) | 0.89 (0.64, 1.24) | 0.88 (0.63, 1.23) |
| IL-12/23 inhibitor | 1.17 (0.44, 3.15) | 1.25 (0.47, 3.33) | 1.23 (0.46, 3.30) | 1.24 (0.46, 3.33) |
| IL-17 inhibitor | 0.37 (0.05, 2.62) | 0.35 (0.05, 2.53) | 0.39 (0.05, 2.75) | 0.39 (0.06, 2.80) |
| IL-6 inhibitor | 0.86 (0.28, 2.66) | 0.75 (0.25, 2.27) | 0.92 (0.30, 2.84) | 0.92 (0.30, 2.81) |
| JAK inhibitor | 1.33 (0.55, 3.20) | 1.19 (0.50, 2.83) | 1.36 (0.57, 3.27) | 1.38 (0.58, 3.31) |
| Rituximab | 2.36 (1.63, 3.41) | 1.92 (1.31, 2.81) | 2.48 (1.72, 3.59) | 2.40 (1.65, 3.50) |
| **COVID-19 hospitalisation** |  |  |  |  |
| Standard therapy | REF | REF | REF | REF |
| Targeted therapy | 1.05 (0.89, 1.24) | 0.99 (0.84, 1.17) | 1.07 (0.90, 1.26) | 1.08 (0.92, 1.28) |
| TNF inhibitor | 0.80 (0.64, 1.01) | 0.79 (0.63, 1.00) | 0.82 (0.65, 1.03) | 0.83 (0.66, 1.05) |
| IL-12/23 inhibitor | 1.29 (0.71, 2.32) | 1.21 (0.67, 2.20) | 1.31 (0.73, 2.37) | 1.35 (0.75, 2.43) |
| IL-17 inhibitor | 1.18 (0.59, 2.37) | 0.98 (0.49, 1.97) | 1.20 (0.60, 2.39) | 1.23 (0.62, 2.46) |
| IL-6 inhibitor | 0.91 (0.41, 2.03) | 0.80 (0.36, 1.79) | 0.93 (0.42, 2.08) | 0.94 (0.42, 2.08) |
| JAK inhibitor | 2.04 (1.23, 3.39) | 1.81 (1.09, 3.01) | 2.05 (1.23, 3.40) | 2.07 (1.25, 3.44) |
| Rituximab | 1.93 (1.41, 2.64) | 1.59 (1.16, 2.18) | 1.94 (1.42, 2.65) | 1.93 (1.41, 2.64) |

Planned comparisons were made between people with IMIDs on any targeted immune modifying therapy (and for each group on specific targeted therapies) compared to people with IMIDs on standard systemic therapy as the reference group.
†Cells with counts less than 5 are redacted to protect anonymity.
**Confounder adjusted:** age, sex, deprivation, smoking status, BMI, specific IMID (inflammatory joint, bowel and skin disease), cardiovascular disease, cancer (excluding non-melanoma skin cancer), stroke, end stage renal failure, chronic liver disease, chronic respiratory disease and diabetes mellitus. **Abbreviations:** HR, hazard ratio; CI, confidence interval; PY, person years; IMID, immune mediated inflammatory disease; BMI, body mass index.

### Text S3. Quantitative bias analysis

#### Methods

We considered severity of immune-mediated inflammatory disease and degree of shielding to be potential confounders which are not available in the data. As such we conducted quantitative bias analysis using E-values to assess how strongly associated these unmeasured confounders would need to be with exposure and outcome to potentially fully explain observed non-null associations (i.e., the association adjusted for both measured covariates and the unmeasured confounder would be null).^21^ In order to apply these methods to the hazard ratio, rather than the risk ratio, we make the assumption that the outcomes are rare.

#### Results

A non-null association was observed between rituximab and COVID-19 death, critical care admission/death, and hospitalisation. For an unmeasured confounder to potentially fully explain the point estimate for the observed confounder-adjusted association the unmeasured confounder would need to be associated with one of exposure or outcome, conditional on measured covariates, by at least risk ratios 2.75, 3.25 and 2.56 for COVID-19 death, critical care admission/death, and hospitalisation respectively (1.46, 1.95, and 1.59 to potentially explain the lower bound of the 95% CI) (**Figures S4-S6, Tables S14-S17**).

Furthermore, to potentially fully explain the point estimate the unmeasured confounder would need to be associated with both exposure and outcome, conditional on measured covariates, by at least risk ratio 1.68, 1.92 and 1.59 for COVID-19 death, critical care admission/death and hospitalisation (1.11, 1.31, and 1.16 to potentially explain the lower bound of the 95% CI).

Additionally, a non-null association was observed between JAK-inhibitors and COVID-19 hospitalisation. To potentially fully explain the confounder-adjusted point estimate (lower bound of 95% CI) the unmeasured confounder would need to be associated with one of exposure or outcome, conditional on measured covariates, by at least risk ratio 3.02 (1.40) and both of exposure and outcome by 1.81 (1.09) (**Figure S7, Table S17).**

#### Discussion

An unmeasured confounder moderately associated with both exposure and outcome could potentially explain the associations of rituximab and JAK inhibitors with COVID-19 outcomes. However, whether this is the case in this study and the extent of true residual confounding depends on both the prevalence of and strength of associations with exposure and outcome of the unmeasured confounders, shielding and severity of IMID, which are not known.

**Figure S4.**
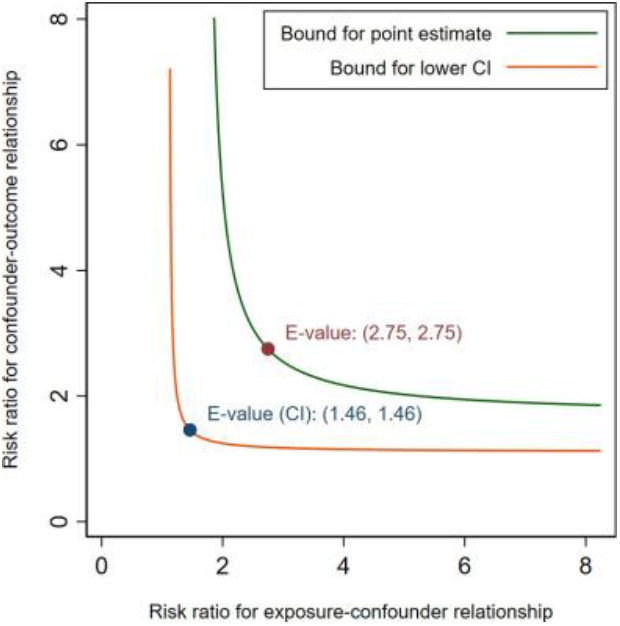
Minimum associations with unmeasured confounder required to potentially explainobserved confounder-adjusted association between rituximab and **COVID-19 death**

**Figure S5.**
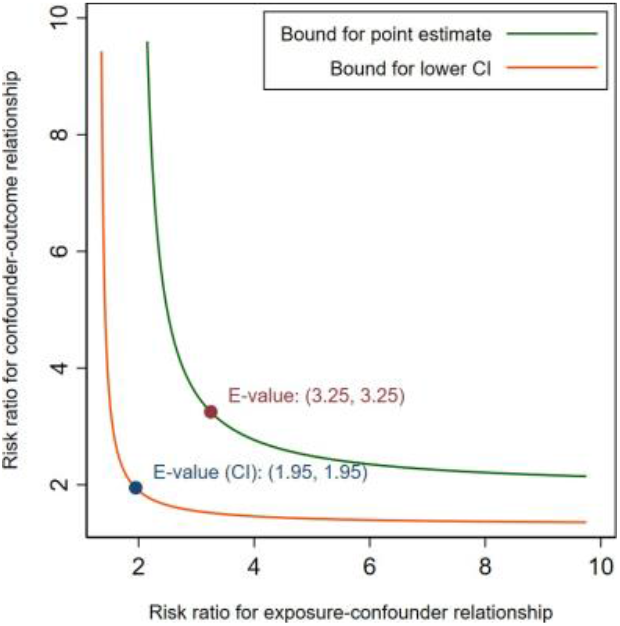
Minimum associations with unmeasured confounder required to potentially explainobserved confounder-adjusted association between rituximab and COVID-19 **critical care admission/death**

**Figure S6.**
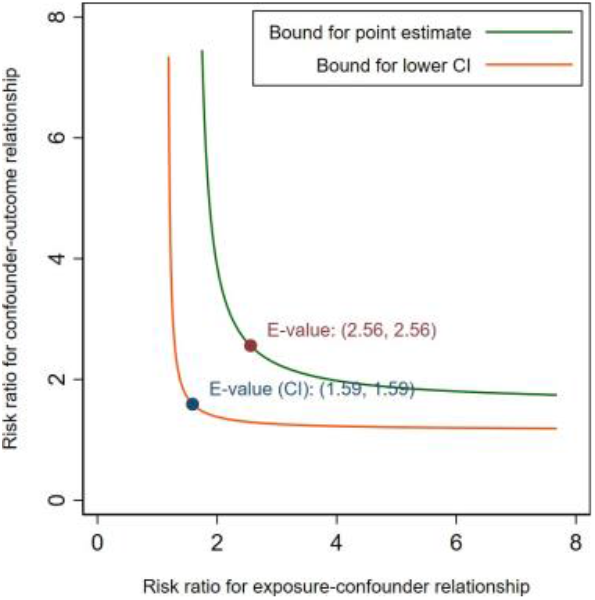
Minimum associations with unmeasured confounder required to potentially explainobserved confounder-adjusted association between rituximab and **COVID-19 hospitalisation**

**Figure S7.**
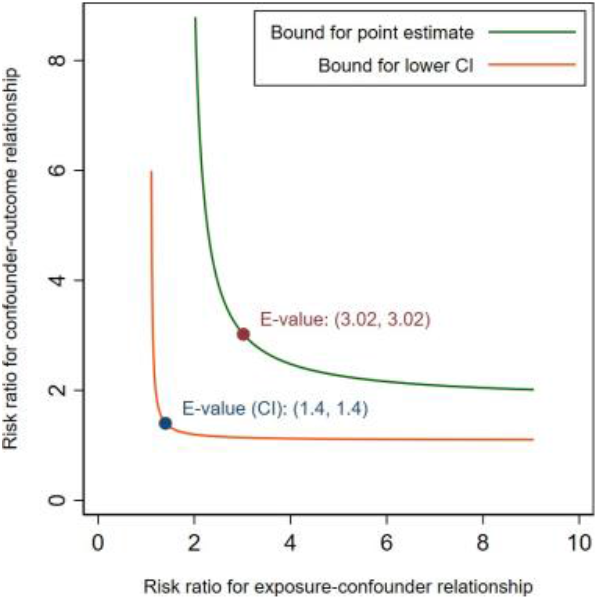
Minimum associations with unmeasured confounder required to potentially explainobserved confounder-adjusted association between JAK inhibitors and **COVID-19 hospitalisation**

**Table S14.** Minimum possible bias-adjusted association between rituximab and COVID-19 death

| Risk ratio<br>between<br>exposure and<br>unmeasured<br>confounder | Risk ratio between unmeasured confounder and outcome |  |  |  |  |
| --- | --- | --- | --- | --- | --- |
|  | 1.2 | 1.5 | 2 | 3 | 5 |
| 1.2 | 1.63 (1.08-2.49) | 1.59 (1.05-2.42) | 1.54 (1.02-2.35) | 1.49 (0.99-2.28) | 1.46 (0.96-2.22) |
| 1.5 | 1.59 (1.05-2.42) | 1.49 (0.99-2.28) | 1.40 (0.93-2.13) | 1.31 (0.86-1.99) | 1.23 (0.81-1.88) |
| 2 | 1.54 (1.02-2.35) | 1.40 (0.93-2.13) | 1.26 (0.83-1.92) | 1.12 (0.74-1.71) | 1.01 (0.67-1.54) |
| 3 | 1.49 (0.99-2.28) | 1.31 (0.86-1.99) | 1.12 (0.74-1.71) | 0.93 (0.62-1.42) | 0.78 (0.52-1.19) |
| 5 | 1.46 (0.96-2.22) | 1.23 (0.81-1.88) | 1.01 (0.67-1.54) | 0.78 (0.52-1.19) | 0.60 (0.40-0.92) |

**Table S15.** Minimum possible bias-adjusted association between rituximab and COVID-19 critical care admission/death

| Risk ratio<br>between<br>exposure and<br>unmeasured<br>confounder | Risk ratio between unmeasured confounder and outcome |  |  |  |  |
| --- | --- | --- | --- | --- | --- |
|  | 1.2 | 1.5 | 2 | 3 | 5 |
| 1.2 | 1.87 (1.27-2.73) | 1.81 (1.24-2.65) | 1.76 (1.20-2.58) | 1.71 (1.16-2.50) | 1.66 (1.14-2.44) |
| 1.5 | 1.81 (1.24-2.65) | 1.71 (1.16-2.50) | 1.60 (1.09-2.34) | 1.49 (1.02-2.19) | 1.41 (0.96-2.06) |
| 2 | 1.76 (1.20-2.58) | 1.60 (1.09-2.34) | 1.44 (0.98-2.11) | 1.28 (0.87-1.87) | 1.15 (0.79-1.69) |
| 3 | 1.71 (1.16-2.50) | 1.49 (1.02-2.19) | 1.28 (0.87-1.87) | 1.07 (0.73-1.56) | 0.90 (0.61-1.31) |
| 5 | 1.66 (1.14-2.44) | 1.41 (0.96-2.06) | 1.15 (0.79-1.69) | 0.90 (0.61-1.31) | 0.69 (0.47-1.01) |

**Table S16.**
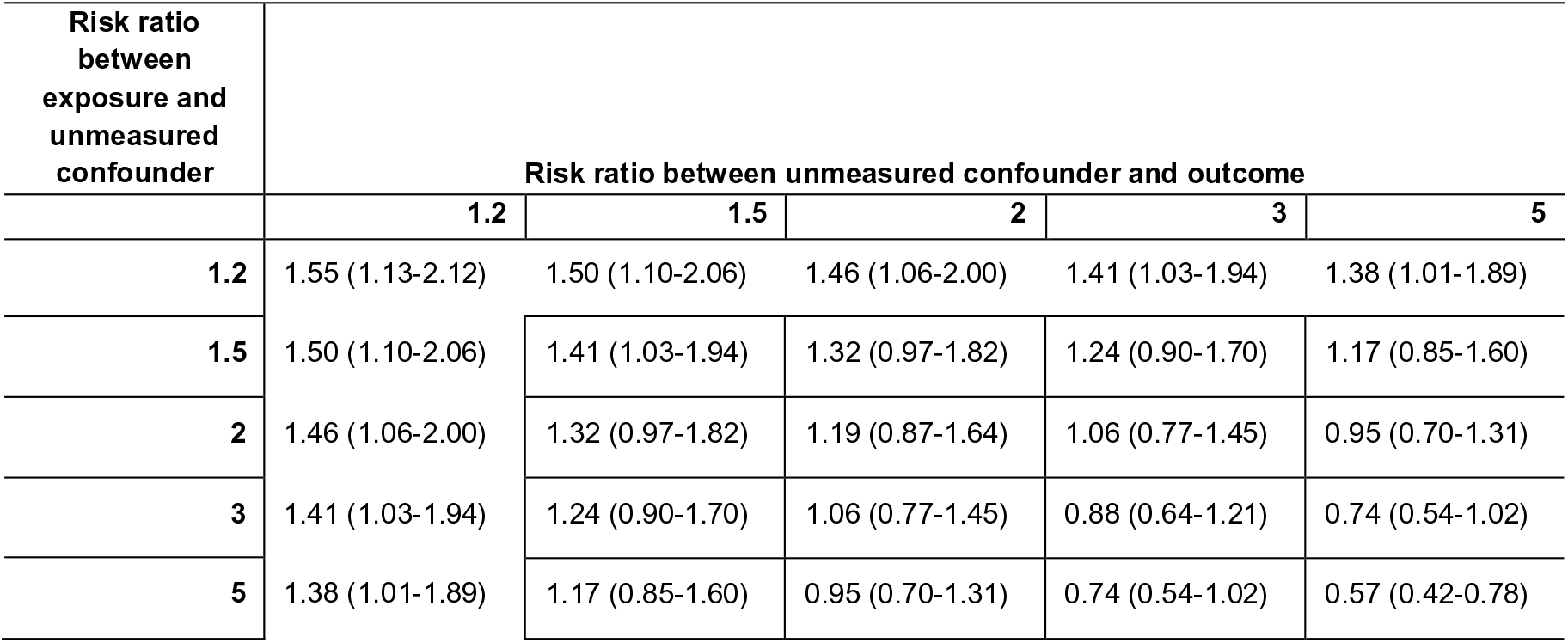
Minimum possible bias-adjusted association between rituximab and COVID-19 hospitalisation

**Table S17.** Minimum possible bias-adjusted association between JAK inhibitors and COVID-19 hospitalisation

| Risk ratio<br>between<br>exposure and<br>unmeasured<br>confounder | Risk ratio between unmeasured confounder and outcome |  |  |  |  |
| --- | --- | --- | --- | --- | --- |
|  | 1.2 | 1.5 | 2 | 3 | 5 |
| 1.2 | 1.76 (1.06-2.93) | 1.71 (1.03-2.84) | 1.66 (1.00-2.76) | 1.61 (0.97-2.68) | 1.57 (0.94-2.61) |
| 1.5 | 1.71 (1.03-2.84) | 1.61 (0.97-2.68) | 1.51 (0.91-2.51) | 1.41 (0.85-2.34) | 1.33 (0.80-2.21) |
| 2 | 1.66 (1.00-2.76) | 1.51 (0.91-2.51) | 1.36 (0.82-2.26) | 1.21 (0.73-2.01) | 1.09 (0.65-1.81) |
| 3 | 1.61 (0.97-2.68) | 1.41 (0.85-2.34) | 1.21 (0.73-2.01) | 1.01 (0.61-1.67) | 0.84 (0.51-1.40) |
| 5 | 1.57 (0.94-2.61) | 1.33 (0.80-2.21) | 1.09 (0.65-1.81) | 0.84 (0.51-1.40) | 0.65 (0.39-1.08) |

